# CovSyn: an agent-based model for synthesizing COVID-19 course of disease and contact tracing data

**DOI:** 10.1101/2025.05.17.25327820

**Authors:** Yu-Heng Wu, Torbjörn E. M. Nordling

## Abstract

The COVID-19 pandemic has demonstrated the shortcomings of epidemiological modelling for guiding policy decisions. Moreover, the modelling efforts resulted in many models yielding different predictions, creating a need to compare these predictions to determine which model is most accurate. We introduce a data synthesis algorithm, CovSyn, designed to generate synthetic COVID-19 datasets providing sufficiently detailed information for benchmarking epidemiological models against a known synthetic ground truth. CovSyn utilizes observed infections with contact tracing, testing, course of disease data, and a contact network based on municipality statistics, which categorises connections into household, school, workplace, healthcare, and municipality. The model’s initial parameters and boundaries are derived from empirical data, including the first community outbreak of COVID-19 in Taiwan and clinical observations. Comprehensive parameter space exploration for optimal results is done by the Firefly algorithm. We demonstrate it and validate our estimates by comparing state transition times, daily social contacts, and associated secondary attack rates against a structured dataset and clinical observations. Our simulations align with prior research and this dataset. Most state transition times from 10,000 simulations are within uncertainty ranges. Daily contact numbers and their distribution across layers match empirical findings. Our model accurately reproduced the first COVID-19 outbreak in Taiwan, achieving high accuracy with observed cumulative confirmed cases (*R*^2^ = 0.9) across daily, 7-day moving average, and 31-day moving average levels. Each synthetic subject contains demographic data (age, gender, occupation), course of disease (latent/incubation periods, testing, isolation, critical illness, recovery, and death dates), and contact network data including daily interactions with infected and uninfected individuals. Our algorithm offers a valid alternative for developing and benchmarking epidemiological models to advance COVID-19 forecasting research.

**Author summary:** We present a novel approach to enhance the testing of epidemiological infection modelling and demonstrate spread prediction accuracy by testing COVID-19 simulation models on Taiwanese synthetic data with known ground truth. CovSyn generates comprehensive synthetic data at the individual level, encompassing demographic characteristics (age, gender, occupation), course of disease (infection dates, symptom onset, recovery dates), and contact tracing information (daily social interactions including household, school, workplace, healthcare, and municipality). It can provide a consistent, standardized synthetic dataset for model evaluation, addressing the previous challenge of comparing COVID-19 models that used disparate data sources and different time periods. In this study, we detail the algorithm and demonstrate its reliability by creating a synthetic dataset for the first outbreak of SARS-CoV-2 in Taiwan and comparing it with the collected Taiwan COVID-19 dataset. Future work will focus on benchmarking state-of-the-art forecasting models using our synthetic data. Through CovSyn’s detailed individual-level data, we aim to advance the development of more accurate epidemiological models.

## Introduction

The COVID-19 pandemic, caused by the Severe Acute Respiratory Syndrome Coronavirus 2 (SARS-CoV-2), has spread worldwide since December 2019. Decision-making processes often rely on mathematical modelling for forecasting and/or intervention comparison. For instance, the UK’s first lockdown was informed by a forecast from Imperial College London. However, this model is highly complex, containing 940 undocumented parameters [1]. It has been shown that the output is highly sensitive to a subset of 19 parameters, leading to results that can vary by up to 300%. Additionally, the model exhibits substantial bias compared to observed data. Rather than calibrating the model to real data, the model relied only on clinical reporting statistics. The Institute for Health Metrics and Evaluation (IHME) model generates forecasts under various scenarios, such as mask usage, vaccination, and antiviral use [2]. Forecasting cumulative death cases with the model in the Global scenario has a 5% error for 4-week forecasts and a 24% error for 12-week forecasts [3]. Furthermore, the peak time accuracy shows a 14-day difference for 4-week forecasts and a 21-week difference for 8-week forecasts. The US Centers for Disease Control and Prevention (CDC) compiled over 80 forecasts from different models to create a single ensemble forecast for public health purposes in the hope of decreasing the uncertainty [4]. Clearly a reliable forecasting model is crucial for guiding disease prevention policies, but already these three examples show that room for improvement exists.

Epidemiological simulation models are designed to predict the spread of an infection in different intervention scenarios. The spread of an infectious disease depends on the virus, the population (demographics, contact networks, and behaviour), the information, and the intervention policy. Thus these models focus on simulating epidemic trends by tracking infection processes within a population or between individuals, including the characteristics, behavior, movement, and disease state transitions of agents, as well as policy [5, 6]. Most COVID-19 models focus on short-term forecasting of disease spread, assessment of the impact of various policies, or a combination of both [7]. Researchers employ diverse approaches, from simple curve fitting to complex machine learning models resulting in different forecasts. For instance, Krymova *et al*. developed a forecasting model based on the underlying trends in observed time series of daily cases and deaths, utilizing robust seasonal trend decomposition techniques [8]. To gain a more comprehensive understanding of the pandemic and predict the consequences of different policies, some researchers model the mechanisms underlying SARS-CoV-2 transmission. These models often adopt the SIR (Susceptible-Infectious-Recovered) framework also known as the compartmental model, incorporating adjustable disease transition parameters. Generally, COVID-19 models can be categorized into two main types: (i) population-based models and (ii) individual- or agent-based models. Each approach offers distinct advantages in simulating and analyzing the complex dynamics of the COVID-19 pandemic.

On the population-based side, the model is either only based on population-level data or the concept of virus spreading at the population level. The daily number of confirmed cases, critically ill, and recovered are the most widely used population data. The IHME model approximates the shape of the epidemic curve of cases and mortality [2]. Their out-of-sample (OOS) predictive performance testing showed their mortality forecasts achieved a median absolute percentage error (MAPE) of 20.2% at 10 weeks of forecasting. They also compared their prediction on surveyed seropositivity to the observed data. Another study proposed a prediction model based on time-dependent SIRVD (Susceptible, Infected, Recovered, Vaccinated, and Deceased) and LSTM (long short-term memory) to forecast the parameters such as transmission rate, recovery rate, and death rate [9]. The input data consists of historical data on confirmed cases, recovered cases, deceased cases, and vaccination data from different countries. While the SIRVD-LSTM model achieved an impressive *R*^2^ of 0.999 for three-day predictions, its accuracy declined to 0.94 for 14-day predictions and further dropped to 0.29 for 28-day predictions.

The individual/agent-based model is based on the interaction network spreading the disease from one individual to another. Imperial College London developed an agent-based stochastic model (CovidSim) and simulated the pandemic under multiple mitigation or suppression policies for the UK and USA, which motivated the UK’s lockdown on March 23, 2020 [10]. Liu *et al*. constructed a contact network with contact layers of households, workplaces, schools, and communities. [11]. Neither model was calibrated using daily or cumulative case data. Instead, it was calibrated using seropositive rate, basic reproduction number (*R*_0_), and other epidemiological parameters. The Institute for Disease Modeling (IDM) developed an agent-based model, COVID-19 Agent-based Simulator (Covasim), simulating each agent’s demographic data, course of disease, and contact network in different social layers [12]. This model has been applied in Africa, Asia-Pacific, Europe, and North America. The model has been calibrated by minimizing the normalized absolute error (NAE) of their prediction to the observed cumulative confirmed cases and deaths, but the achieved error is not reported. Unlike population-based models, agent-based models are typically not validated directly by standard metrics such as MAE, but rather by aiming to be consistent with clinical observations.

Current epidemiological forecasting models are calibrated based on population data. This requires precise population measurements such as the accurate daily number of infections or deaths, which have been shown to be biased [13–18]. Reliable forecasting was impossible at the beginning of the COVID-19 pandemic due to the lack of reliable public data [19, 20]. Individual-level data is more informative from a modelling perspective but is almost completely lacking in public databases, limiting the development of epidemiological models. We have thus collected a comprehensive structured individual-level Taiwan COVID-19 dataset [21] and calibrated our data synthesis model on it.

Epidemiological models have only in a few cases been benchmarked. At the beginning of the COVID-19 pandemic, Friedman *et al*. compared 7 models’ forecasting accuracy for 1-12 week daily cases using MAPE and peak time prediction. Their findings showed that the SIK-Jalpha and IHME models performed best [3]. However, the 12-week forecasts still yielded 24% MAPE. The CDC COVID-19 Forecast Hub consolidated forecasts from multiple teams for US daily confirmed cases, hospitalizations, and deaths. They evaluated results using mean absolute error (MAE) and weighted interval scores (WIS), maintaining updated model comparisons on their website until June 23, 2023 [4]. Subsequently, Chharia *et al*. analyzed these models’ predictive capabilities [22], revealing that two-thirds underperformed the CovidHub-Baseline model (where the median prediction at all future horizons is the most recent observed value) and one-third fell short of linear trend forecasts (which use extrapolation based on the slope of change in reported active cases from the two weeks preceding the forecast date). Notably, these benchmarking efforts relied solely on comparing forecasting results provided by research teams against subsequently reported daily cases, which seropositive studies have shown to be underreported [13–16]. None conducted direct model performance comparisons using standardized data with known ground truth. In the field of Systems Biology, *e*.*g*. the DREAM network inference competition utilized in silico (synthetic) data as ground truth to evaluate the performance of network inference methods [23]. Specifically, this approach was implemented in the DREAM 2, DREAM 3, and DREAM 4 in silico network challenges. Recently, a comprehensive benchmark of deep time series models has been conducted and suitable structures for distinct tasks have been identified [24]. Following this idea, we believe a fair comparison of COVID-19 models would require a synthetic dataset where all quantities are known.

Moreover, synthetic epidemiological data remains underutilized in the field. As a notable exception, a Taiwanese research team developed a compartmental model of hepatitis A virus (HAV) transmission structured according to human immunodeficiency virus (HIV) risk status. By fitting this model to the actual epidemic curve and incorporating parameters from literature and their case-control study, they simulated the counterfactual scenario without vaccination—projecting 7,153 cases compared to the observed 1,352 cases [25]. For COVID-19, we identified only a study that employed simple stochastic agent-based models with SIR, SEIR, and SEAIR structures to evaluate six different *R*_0_ estimation methods.

The variety of model structures and specific input data used in COVID-19 modelling, makes it impractical to benchmark their performance through direct comparison. To address this issue, we here propose an algorithm for generating COVID-19 synthetic data that is comprehensive enough to cover most of the input data requirements of state-of-the-art epidemiological forecasting models—both for initial settings and for calibrations—thereby enabling fair performance comparisons in the future.

In the following Results section, we first present an overview of CovSyn and data followed by an example of commonly used epidemiological data required for implementing COVID-19 models. Then, we demonstrate what variables CovSyn can generate, how these variables are translated to the commonly used variables in COVID-19 models, and how closely the synthetic data’s important epidemiological statistics match clinical observations of COVID-19. Next, we introduce the optimization process and show that our Firefly optimization has successfully guided the system to an optimal solution. Finally, we demonstrate how closely our synthetic data matches the structured Taiwan COVID-19 dataset by comparing Kaplan-Meier plots, comparing the synthetic data with Taiwanese clinical observations on social contacts and secondary attack rates, and comparing the synthetic data with the first COVID-19 outbreak in Taiwan. In the Discussion section, we discuss the results and limitations of our work. In the Materials and Methods, we present CovSyn, its implementation, and our experiments.

## Results

CovSyn is an agent-based model designed to generate synthetic COVID-19 data at an individual level implemented in Python 3.10. This enables benchmarking and development of epidemiological models through access to a ground truth dataset implemented.

The framework operates on two distinct input data streams: initialization data and training data. Initialization data provides baseline parameters required for simulation setup. Training data facilitates model calibration and refinement to ensure predictive accuracy. When CovSyn is used without model optimization and training data, it attempts to load optimized model parameters from a previous optimization. If this attempt fails, it resorts to default values and boundaries defined in `parameters_for_training.py`.

A key aspect of CovSyn is its utilization of the individual-level Taiwan COVID-19 dataset, encompassing 578 subjects [21]. Unlike the commonly used aggregate daily case counts, CovSyn incorporates detailed individual records. The overall workflow of CovSyn is illustrated in Fig 1. The demographic data and epidemiological parameters serve as inputs for constructing disease state transition, demographic distributions, clinical statistics, household, school, workplace, hospital, and municipality data. For the demographic data, the region of interest is defined as the country, county, city, or town that one wants to simulate the pandemic for. Demographic data are collected from the National Development Council (NDC), Ministry of Interior (MOI), Ministry of Education (MOE), Executive Yuan (EY), and Ministry of Health and Welfare (MOHW). Sources for these datasets can be found in the Demographic and social dataset section. Epidemiological parameters come from various literature sources, with full explanations provided in the Parameter setting section.

**Fig 1.**
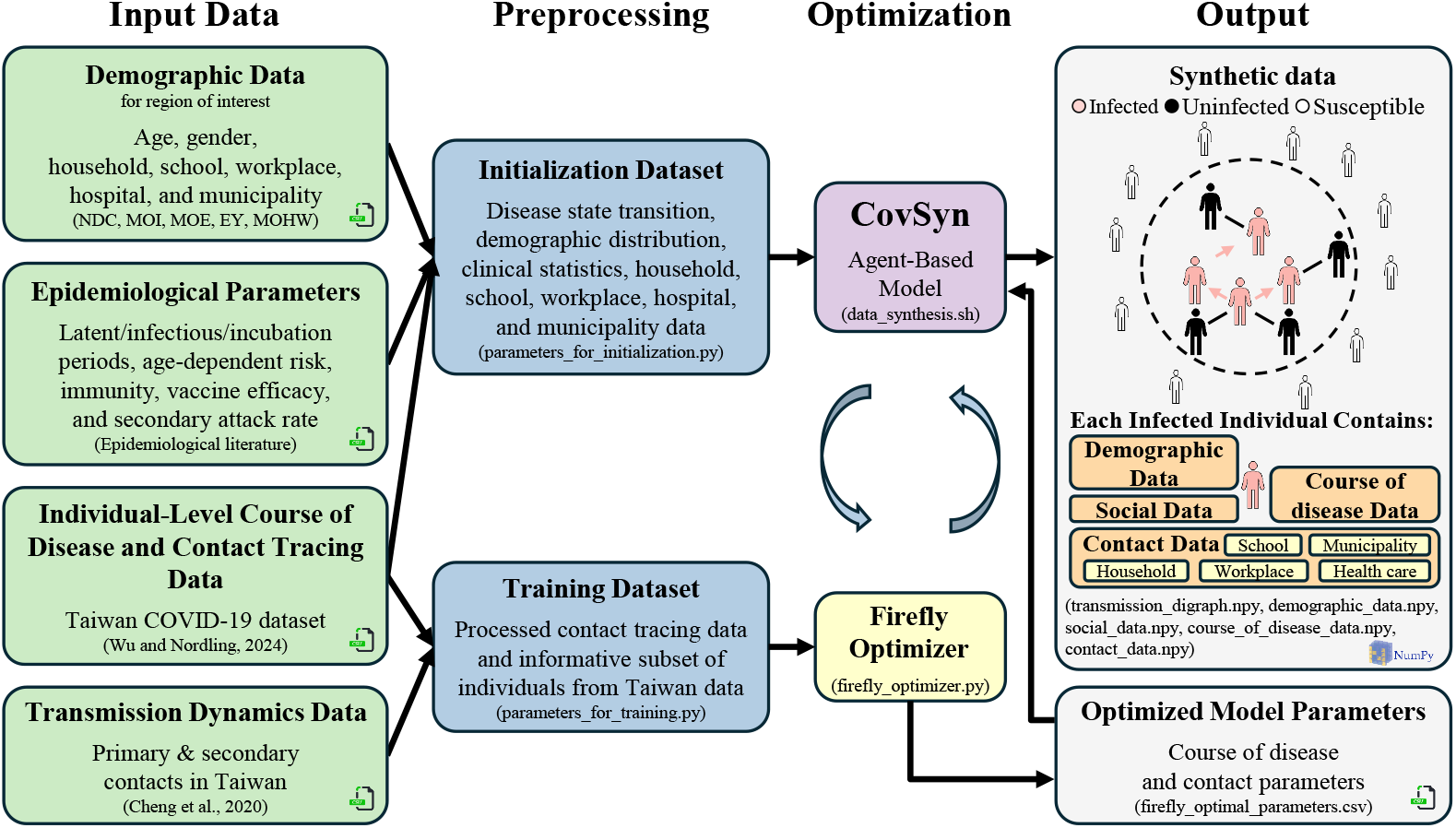
Schematic representation of the CovSyn framework for generating synthetic COVID-19 data. The pipeline processes input data in CSV format into an initialization dataset for model setup and a training dataset for optimization. CovSyn trains using an informative subset of Taiwan COVID-19 dataset [21] alongside transmission dynamics data [26] and optimizes iteratively using the Firefly algorithm. The model outputs synthetic individual-level course of disease and contact information data, capturing all infected individuals with their infected and uninfected contacts categorized into five different layers: household, school, workplace, health care, and municipality. Each infected individual retains demographic, social, course of disease, and contact data, facilitating epidemiological modelling and analysis.

The Taiwan COVID-19 dataset contributes to both the initialization and training phases. Disease state transition distributions are extracted and incorporated into the initialization dataset, while a curated subset of the Taiwan dataset is used for model training. Due to data sparsity, subjects with the most complete feature sets, those with at least four recorded features, were selected. This resulted in a training dataset with dimensions 121 *×* 9. A detailed methodology for data selection and processing is provided in the Materials and methods section. Additionally, transmission dynamics data from Cheng *et al*. [26] were integrated into the training dataset. Python files `parameters_for_initialization.py` and `parameters_for_training.py` handle the data preprocessing. The Firefly optimization is implemented in `firefly optimizer.py` and the bash script `data_synthesis.sh` executes the CovSyn agent-based model simulation. The optimized parameters are shown in Table S2 to Table S9 and stored in a CSV file `firefly_optimal_parameters.csv`.

CovSyn generates a contact network that includes both infected and uninfected individuals. The data for each infected individual is stored, including distinct demographic, social, course of disease, and contact information, which can be readily accessed for analysis. NumPy files (`.npy`) store the structured output data.

### Synthetic variables and commonly used epidemiological data

CovSyn generates data for each individual across four main categories: demographic, social, course of disease, and contact tracing. In brief, the demographic data includes age, gender, and job; the social data contains information about how many people the individual can potentially contact in different layers; the course of disease data includes the disease state transition data; and contact data includes the daily contact data in different layers. The detailed variables that CovSyn can generate are illustrated in the top plot of Fig 2. The visualization uses orange nodes to represent the four main categories, each connected to their corresponding detailed variables (shown in red) that CovSyn directly generates.

**Fig 2.**
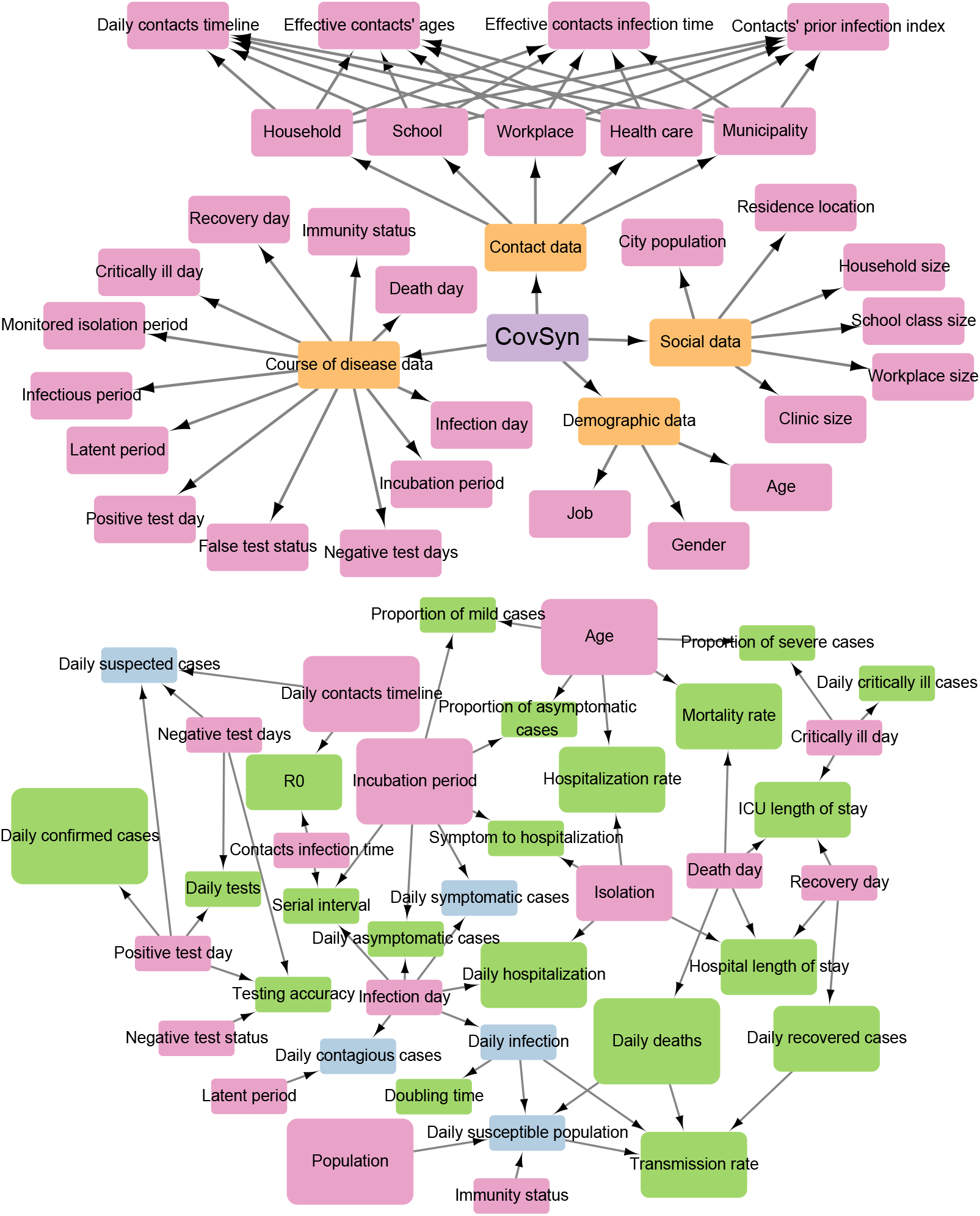
Visualization of CovSyn-generated variables (top plot) and their relationship to common epidemiological model variables (bottom plot). Orange nodes indicate the four main data categories and red nodes show variables directly generated by CovSyn. In the bottom plot, the green and blue nodes indicate derived variables that can be easily calculated from the connected red nodes. Green nodes represent variables commonly used in epidemiological models listed in Table 1. Blue nodes indicate supplementary epidemiological derived variables not listed in Table 1 but potentially valuable for modelling. Node size is proportional to the frequency of use in modelling, as documented in Table 1.

To demonstrate the practical applications of CovSyn-generated data in implementing COVID-19 models, we first identify initialization and training data requirements for both general model structures and some state-of-the-art models. Table 1 outlines potential models that could be trained and/or tested using our synthetic data. Inspired by the model purpose classifications proposed by Willem *et al*. [5], we define three primary purposes for COVID-19 models. First, ‘explanation’ models aim to elucidate transmission dynamics and examine the effects of various model assumptions and parameter values on the outcomes. Second, ‘intervention’ models are designed to assess diverse mitigation measures, thereby providing evidence-based insights to policymakers through methodologies grounded in our current understanding of viral transmission dynamics. Finally, ‘forecast’ models endeavor to predict future epidemic trends by leveraging historical data and employing statistical analyses.

**Table 1.**
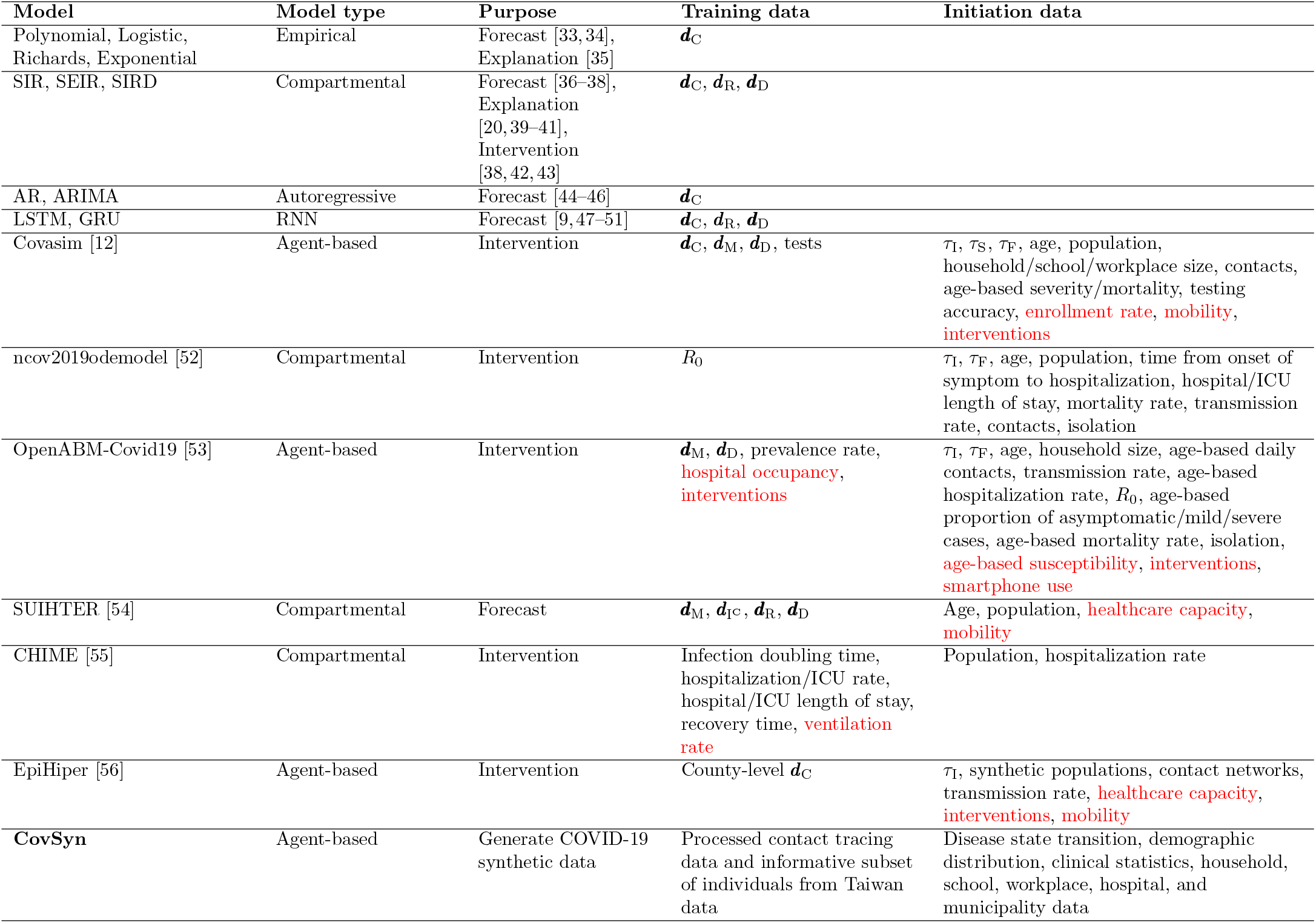
Comparison of COVID-19 models based on model type, purpose, required training data, and initiation data. Mathematical symbols: ***d***_C_ daily confirmed cases, ***d***_M_ daily hospitalizations, ***d***_I_C daily critically ill cases, ***d***_R_ daily recovered cases, ***d***_D_ daily deaths, *τ*_I_ incubation period, *τ*_S_ serial interval, and *τ*_F_ infectious period. Data not included in our synthetic dataset is highlighted in red.

It is worth noting that we do not cover risk prediction models. These models are designed to predict the likelihood of specific outcomes based on various risk factors and data inputs. In the context of COVID-19, they are employed for patient prognosis, identifying individuals in the general population at increased risk of infection, and predicting the likelihood of hospital admission or death due to the disease. However, they do not include disease transmission dynamics. Thus, strictly speaking, they do not fall under our definition of COVID-19 simulation models. Well-known models all require detailed individual data such as demographics, medical conditions, and clinical assessments like vital signs and comorbidities [27–32]. This type of data is not covered in our study as it is typically not openly available.

To further demonstrate how CovSyn data maps to the commonly used data listed in Table 1, we present a directed graph visualization at the bottom of Fig 2. Red nodes, consistent with the top plot, represent variable directly generated by CovSyn, which connect to commonly used epidemiological variable (shown in green). The visualization shows how different sources of CovSyn data combine to generate a specific epidemiological variable by following the directional arrows. For example, calculating daily symptomatic cases requires both infection day and incubation period for each individual. Moreover, nodes in blue represent supplementary variable not listed in Table 1 but potentially valuable for epidemiological modelling. One example is the daily infection data, which, while difficult to measure in practice due to the extensive contact tracing and widespread testing required, can be valuable for implementing COVID-19 models. We provide code that from our synthetic data calculates these common epidemiological variable in `example_data_mapping.py`.

We conducted 100 Monte-Carlo simulations with a simulation time limit of 365 days, to evaluate the distribution of mapped statistics, generating a total of 3,176 infected individuals. We calculated key epidemiological metrics including the duration of disease stages and transmission intervals, comparing them with reported values from various sources. Our analysis yielded the mean days for the latent period (3.8), incubation period (5.1), infectious period (21.8), generation time (8.2), and serial interval (8.2), respectively. The distributions of these metrics are visualized in Fig 3 and Table 2. Our estimations for the infectious period, generation time, and serial interval tend to be higher than reported mean values, though all fall within the reported 95% confidence interval ranges. It’s worth noting that latent periods and generation times are less frequently reported in studies due to the inherent difficulty in determining precise infection dates. We estimate the *R*_0_ by calculating the average number of secondary cases per index case.

**Table 2.** Epidemiological parameters of COVID-19 from literature and CovSyn model estimates. Literature CI Range represents the combined span from the minimum lower bound to the maximum upper bound of 95% confidence intervals reported across all cited studies.

| Parameter | Literature Mean | Literature CI Range | CovSyn Mean Estimate | References |
| --- | --- | --- | --- | --- |
| Latent period (days) | 4.1–5.5 | 0.1–27.8 | 3.8 | [26, 57] |
| Incubation period (days) | 5.2–8 | 0.4–15.8 | 5.1 | [26, 58, 59] |
| Infectious period (days) | 3.5–20 | 0–24 | 21.8 | [60] |
| Generation time (days) | 3–5.2 | 0.4–11.5 | 8.2 | [59, 61] |
| Serial interval (days) | 3.0–7.6 | -4.5–19 | 8.2 | [61–63] |
| $R_0$ | 0.3–6.7 | 0–12.3 | 0.4 | [64–72] |

**Fig 3.**
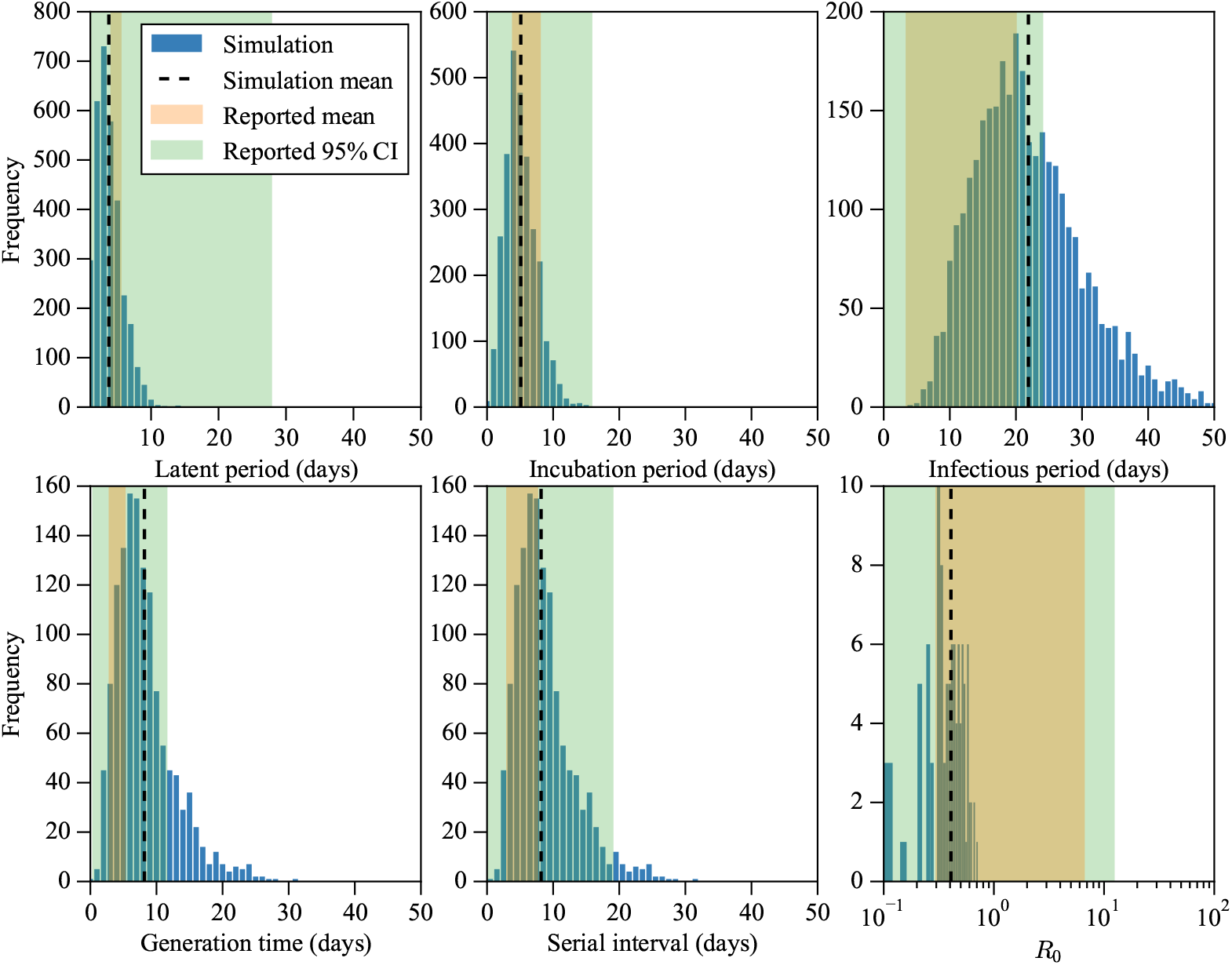
Statistics of the synthetic data generated by CovSyn. The latent, incubation, and infectious periods, as well as generation time and serial interval, are directly sampled from distributions in our model. The basic reproduction number (*R*_0_) is calculated as the average number of effective contacts per infected case. Blue histograms represent simulation results with black dashed lines indicating mean values. The orange shaded areas show the range of mean values reported across multiple studies listed in Table 2, while green shaded areas represent the span from the minimum lower bound to the maximum upper bound across all confidence intervals reported in the literature.

### Parameter setting and optimization results

The CovSyn parameters are divided into demographic, course of disease, and social contact parameters. The demographic parameters consist of 25,505 fixed values. The trained parameters include the course of disease parameters containing 161 trainable parameters and social contact parameters consisting of 37 trainable parameters. The detailed parameter settings are shown in the Epidemiological parameters section.

The fireflies optimization process achieved successful convergence, as illustrated in the top two subplots of Fig 4. The visualization traces the movement of all fireflies from their initial positions (red dots), through their worst performance points (black crosses) and best performance points (black triangles), to their final positions (blue dots). The final positions cluster closely around an optimal solution, demonstrating effective convergence of the algorithm. The heatmap shows the Euclidean distance between each final firefly, revealing the formation of one large cluster and two smaller clusters. We also show the convergence of costs for each layer including household, healthcare, other, energy, and total cost. The middle four subplots show the convergence of costs for the four layers and the bottom two subplots show the Energy distance between the Taiwan matrix which is used in the training process (121 *×* 9, included in the training process) and the full Taiwan matrix (578 *×* 9). Both distances converge to smaller values, indicating that the optimization process was successful.

**Fig 4.**
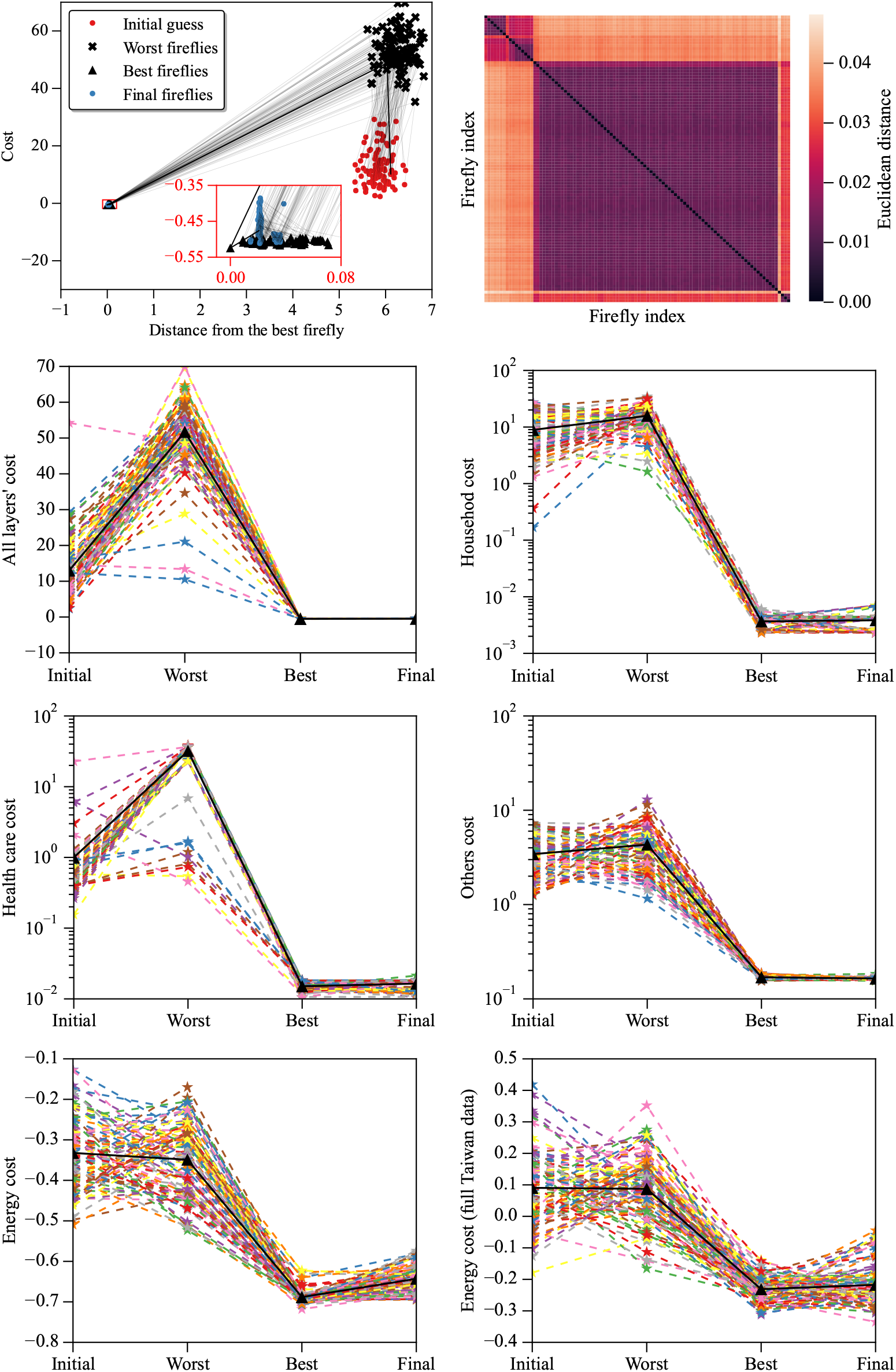
Convergence of each layer’s cost including household, health care, other, energy and total cost. The top left plot indicates that the system has multiple local minima. We saved each Firefly’s initial guess, worst cost, best cost, and final cost. The red rectangle shows the zoomed-in plot around the final fireflies. The top right plot is the heatmap of the distance matrix of the final fireflies. The coloured lines with start markers are the 100 fireflies. The black line with triangle markers is the average of the 100 fireflies.

Firefly optimization has resulted in an energy distance of -0.21 between our synthetic dataset and the Taiwan COVID-19 dataset, with a p-value of 0.99, meaning that we cannot reject the null hypothesis that both datasets are from the same distribution [73, 74].

### Validation

We validate CovSyn by evaluating its ability to reproduce both individual-level and population-level COVID-19 data. We implemented three scenario Monte Carlo simulations to evaluate the model’s performance in generating course of disease, contact data for each layer, and daily local confirmed cases as well as deaths. The detailed setting of each scenario is described in the Monte Carlo simulations section.

Our algorithm successfully mimics statistical observations while producing detailed epidemiological progressions. Fig 5 demonstrates that the synthetic course of disease data is close to the observed Taiwan COVID-19 dataset, and Fig 6 illustrates that our synthetic contact tracing data exhibit the same contact patterns across all layers. All layers had higher contact probabilities before symptom onset except for the healthcare layer. Fig 7 demonstrates the ability of CovSyn to capture the patterns in Taiwanese population-level data.

**Fig 5.**
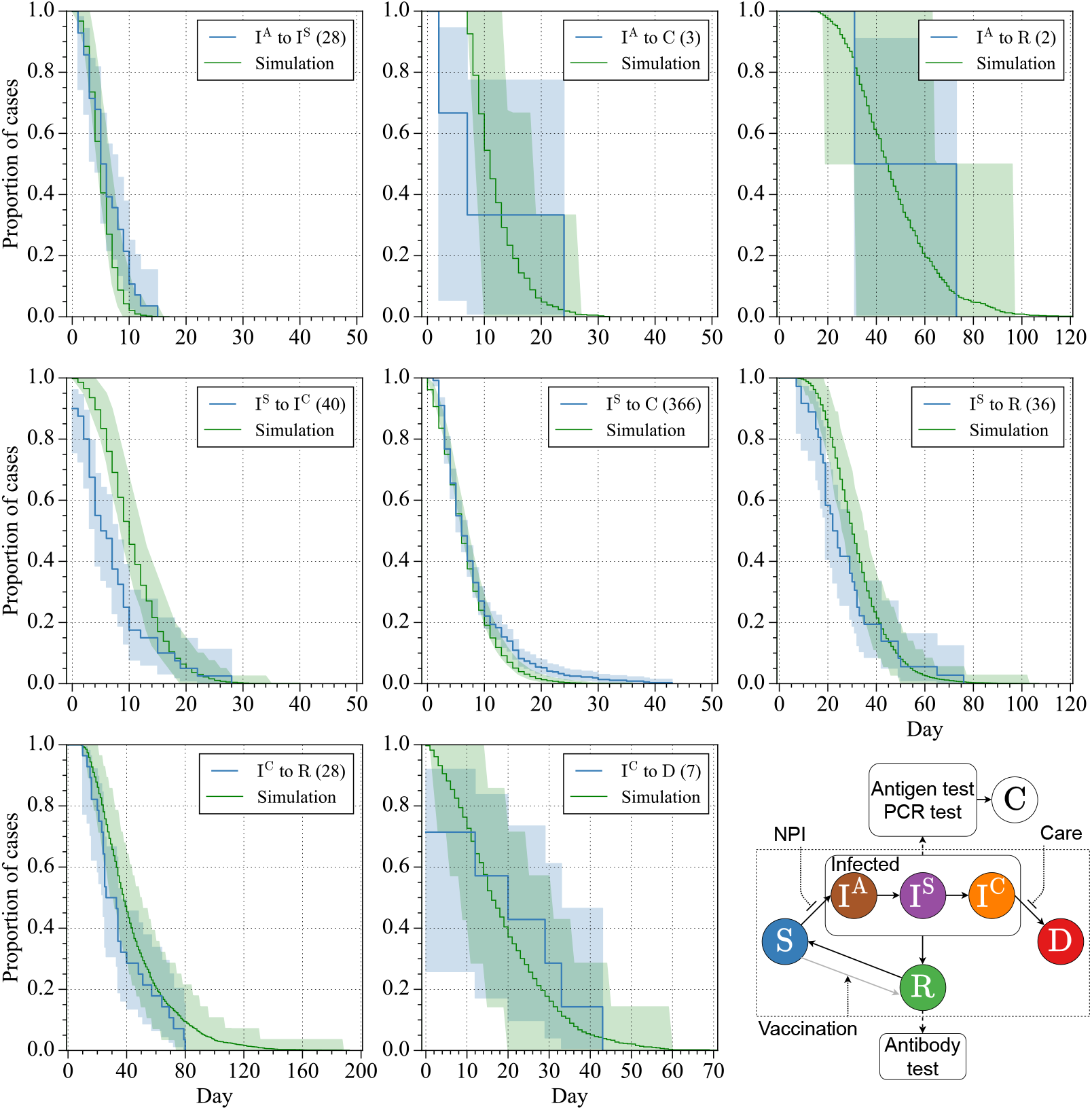
State transition verification through Kaplan-Meier plot. The blue lines are extracted from the Taiwan COVID-19 dataset and the green lines are based on 10,000 simulations. The uncertainty bound of the simulation (green shaded area) is calculated by bootstrap and the uncertainty bound of the data (blue shaded area) is calculated by the Greenwood’s formula. The different states are denoted as asymptomatic (I^A^), symptomatic (I^S^), critically ill (I^C^), confirmed (C), recovered (R), and death (D). The bottom right plot is the state transition network with detailed infection state descriptions. The probabilities of state transitions are affected by inhibitor nonpharmaceutical interventions (NPI) and intensive care units (Care) and activator vaccination. In practice, the infection state can be measured by antigen test and PCR test. The recovered/immune state can be measured by an antibody test.

**Fig 6.**
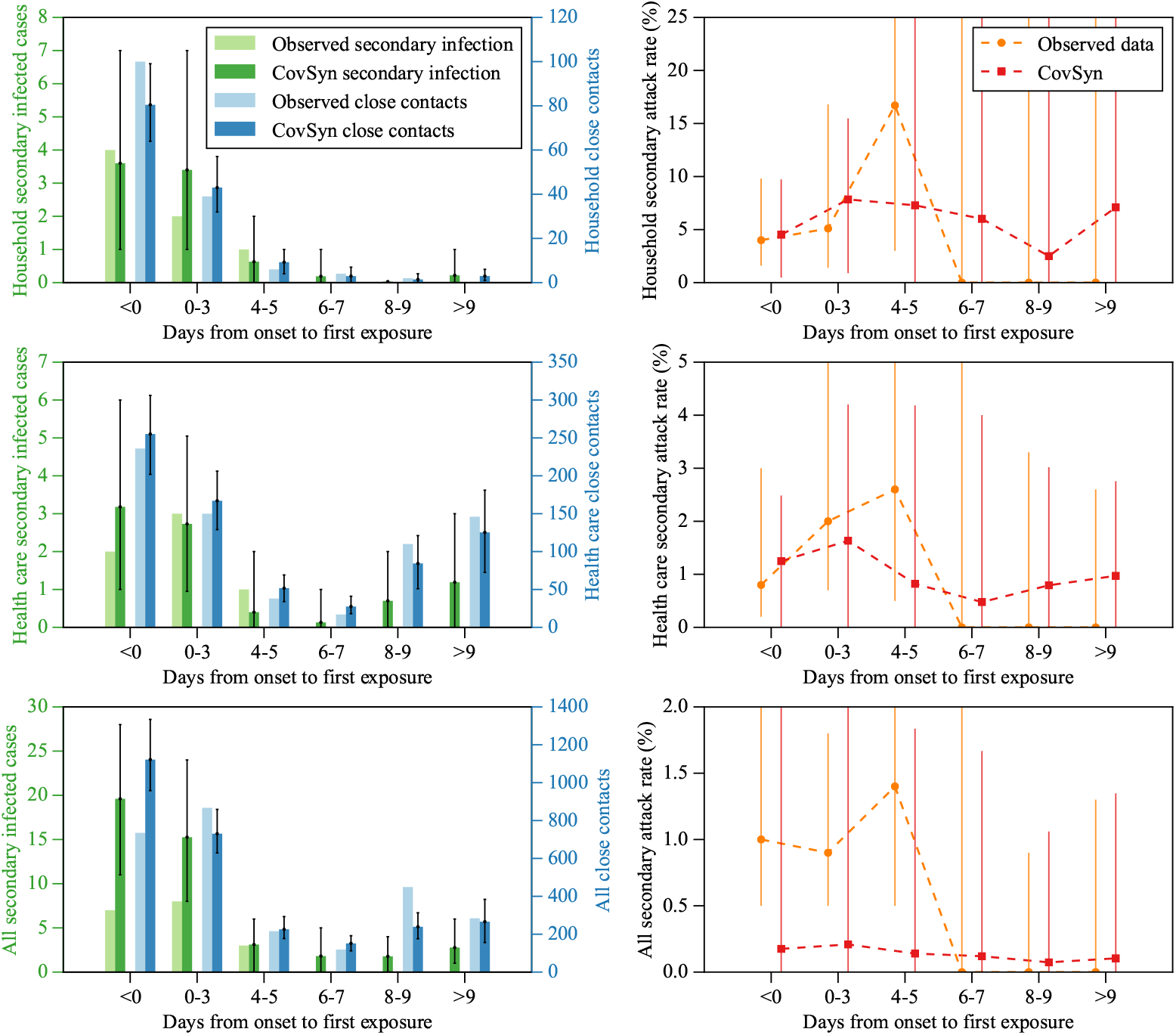
Comparison between CovSyn simulated data and observations from Cheng et al. 2020 across different contact layers: household (top), health care (middle), and all contacts (bottom). Left panels show secondary infected cases and close contacts. Right panels display the secondary attack rate, defined as the ratio of secondary infected cases to close contacts, expressed as a percentage. CovSyn was run for 100 Monte Carlo simulations, each with 100 source cases, to generate the 95% confidence intervals. The comparison demonstrates that our synthetic data exhibit the same trend in each layer.

**Fig 7.**
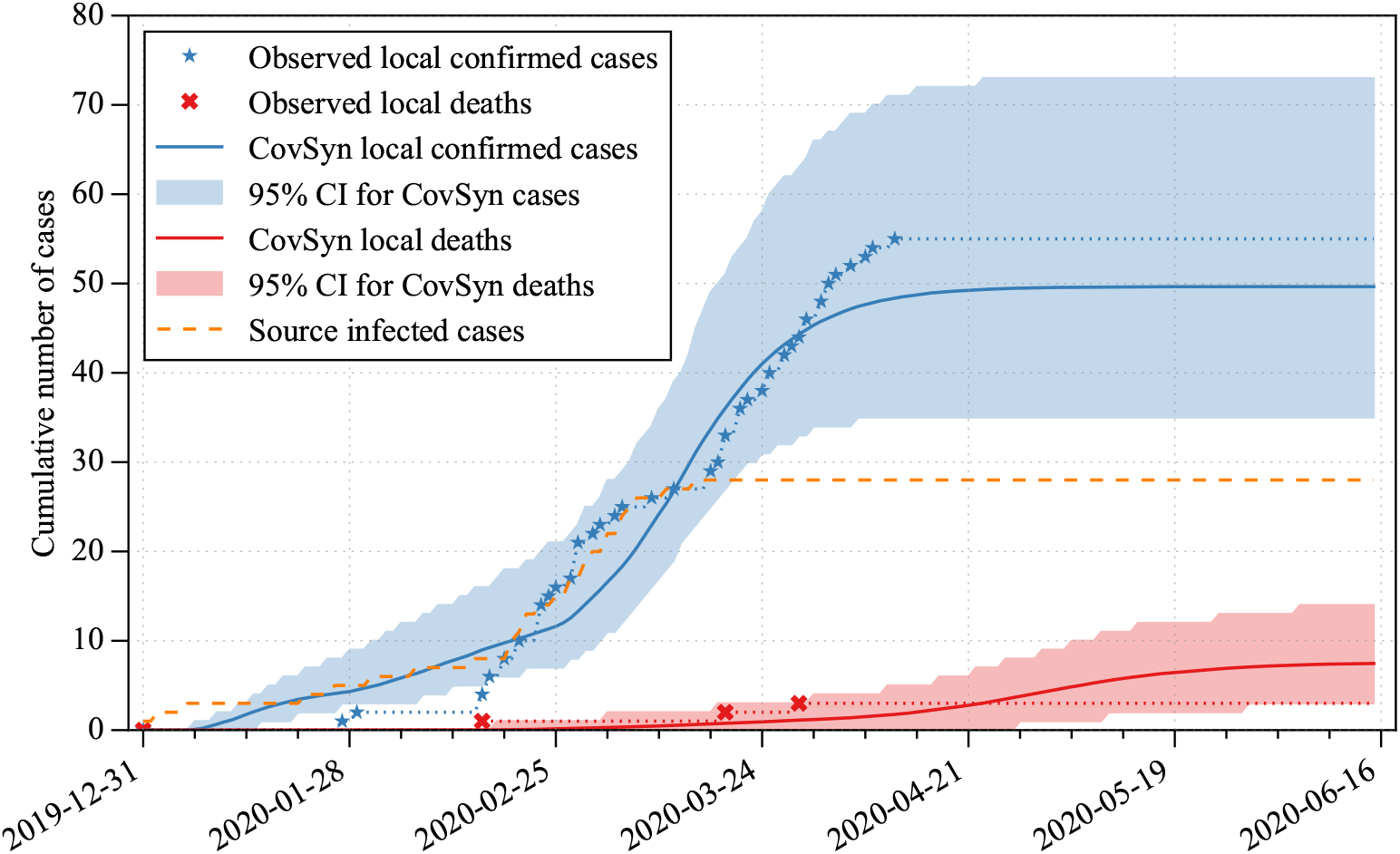
Comparison of observed Taiwan COVID-19 population-level data with CovSyn simulations. Blue stars and red dots show observed local cases and deaths, respectively. Dashed orange line indicates the 28 seed infections. Blue/red solid lines represent mean predictions from 1,000 Monte Carlo simulations, with 95% confidence intervals shown as shaded areas.

### Validation of individual-level data

We compared the synthetic data generated as described in Section Course of disease simulation with the Taiwan COVID-19 dataset shown in Fig 5. Most of the simulation results stay close to the 95% confidence interval of the Taiwanese data. Some of the transition times are slightly longer than those in the Taiwanese data, including symptomatic to critically ill, symptomatic to recovered, and critically ill to recovered.

Cheng *et al*. investigated the close contacts of 100 symptomatic index subjects and 2,670 close contacts in Taiwan [26]. The second scenario of the Monte Carlo simulation was implemented to reproduce their results, with detailed settings described in the Secondary close contact simulation section. The contact type, such as household, family, health care, and others and their contact number versus the secondary attack rate was studied. We reproduced the bar chart of the layers of household, health care, and all contacts. The contacts of the 100 source cases were generated, resulting in 2491 close contacts, and the result is shown in Fig 6. In this figure, we focus on the difference between the two secondary attack rates. For a more comprehensive view covering the entire uncertainty range, readers can refer to Fig S1. Since the study was conducted at the beginning of the pandemic, we set the vaccination rate to 0. In order to reflect the phenomenon that the first exposure number increases after day 8 after symptom onset for the health care layer, we implement the weighting function for the contact probability.

Cheng *et al*. analysed Taiwanese COVID-19 data from 2020-01-15 to 2020-03-18, which covered half of the duration of the first outbreak in Taiwan (2020-02-16 to 2020-04-11) [26]. However, in their methodology, they only reported the initial day of contact between individuals. For example, if a close contact first interacted with the index case before symptom onset (day 0), they were only counted in the ‘zero days’ category, regardless of any subsequent interactions. This means that even if someone lived continuously with the index case from day 0 until isolation, they were only counted as having contact on the first day. This methodological choice likely resulted in overestimated contact and secondary attack rates for the early days of infection, while potentially underestimating these rates for later days.

In contrast to the reproduced results, we also show the detailed synthetic contact days instead of merely the first exposure date. The bar chart for each layer is shown in Fig S5. This figure shows the details of daily contact with their contact layer.

### Validation of population-level data

To assess the ability of CovSyn to capture the patterns in Taiwanese population-level data, we compared CovSyn-generated 1,000 simulations with Taiwanese population-level COVID-19 data with the performance metrics in Table 3 and results shown in Fig 7.

**Table 3.** Goodness of Fit (GOF) metrics comparing Taiwanese population-level COVID-19 data with CovSyn synthetic data outputs. The metrics include Root Mean Squared Error (RMSE) and coefficient of determination (*R*^2^). Subscript c denotes cumulative values. Centered Moving Average (MA) over stated period used. All metrics are presented with 95% confidence intervals.

|  | **RMSE** | **RMSE<sub>c</sub>** | **$R^2$** | **$R^2_c$** |
| --- | --- | --- | --- | --- |
| Daily Confirmed | 0.9<br>(0.8, 1.0) | 7.6<br>(3.1, 15.8) | -0.4<br>(-1.0, -0.1) | 0.9<br>(0.5, 1.0) |
| Daily Deaths | 0.3<br>(0.2, 0.4) | 2.3<br>(0.9, 4.8) | -2.7<br>(-5.8, -1.0) | -2.8<br>(-12.9, 0.6) |
| 7-day MA Confirmed | 0.5<br>(0.4, 0.6) | 7.5<br>(2.9, 15.8) | 0.1<br>(-0.4, 0.5) | 0.9<br>(0.5, 1.0) |
| 7-day MA Deaths | 0.1<br>(0.1, 0.2) | 2.3<br>(0.8, 4.8) | -4.5<br>(-11.2, -0.9) | -2.8<br>(-13.0, 0.6) |
| 31-day MA Confirmed | 0.2<br>(0.1, 0.3) | 6.9<br>(1.8, 15.6) | 0.7<br>(0.4, 0.9) | 0.9<br>(0.5, 1.0) |
| 31-day MA Deaths | 0.1<br>(0.0, 0.1) | 2.2<br>(0.7, 4.7) | -8.8<br>(-29.5, -0.7) | -2.7<br>(-13.0, 0.7) |

The Goodness of Fit (GOF) metrics presented are the average values calculated across all 1,000 Monte Carlo simulations. The full version of the performance metrics table is provided in Table S11.

A critical limitation in our validation approach was the absence of actual infection dates in available data, necessitating our manual identification of the source infection dates through analysis of infection chains in contact tracing records. To initialize our simulations, we identified 28 cases as seed infections from available contact tracing data. These seed infections initiate the branching process in our model and later become part of the confirmed case counts at their respective confirmation dates, which are determined through the stochastic simulation process. The detailed methodology is provided in the Taiwan first outbreak section.

## Discussion

The epidemiological parameters estimated by our model generally align with values reported in the literature as shown in Fig 3 and Table 2. Our 21.8 day estimate of the infectious period is near reported, likely due to constraints in our algorithm requiring the isolation date to fall after the end of the latent period and before the end of infectious period (Fig 9). This may create selection pressure toward longer infectious periods, which consequently could affect generation time and serial interval estimates due to their inherent linkage to infectious period duration. In future work we intend to test relaxing this constraint. Our estimation of *R*_0_ is 0.4 which is relatively low. This value results from our model’s calibration to the clinical research of Cheng *et al*. [26], who early in the pandemic, conducted a study examining 100 source cases’ transmission that identified only 22 paired index-secondary cases due to successful contact tracing and isolation, as well as reduced social interactions in Taiwan. The report of Cheng *et al*. corresponds to *R*_0_ *≈* 0.22, which is closely matching our estimate.

Kaplan-Meier plots (Fig 5) show that most of the simulation results stay within the 95% confidence interval of the Taiwanese data. Area between curves analysis (Table S10) reveals bigger divergence for the I^S^ to I^C^ (3.5), I^S^ to C (-1.14), I^S^ to R (4.75), and I^C^ to R (9.45). These temporal shifts associated with the transition from the symptomatic state I^S^ are likely attributable to clinical reporting practices, where symptom onset dates are often retrospectively adjusted following hospitalization, creating potential reporting bias in the reference dataset. Additionally, the transition I^C^ to R fits well initially but has a much longer tail after day 80, causing the area between curves to be larger. Our model predict longer recovery times, in particular for the 5% subjects in the tail. Importantly, these transition time discrepancies have not yet been incorporated into our objective function for optimization. Future iterations of the CovSyn algorithm will address this issue by integrating these curve-based metrics directly into the optimization process.

We compared secondary infections, close contact numbers, and resulting secondary attack rates from our synthetic data with observations from Cheng *et al*., 2020 [26] (Fig 6). CovSyn successfully captures similar trends across all layers, with particularly accurate replication in the healthcare layer, which exhibits a distinctive pattern of increased close contacts after day 8 post-symptom onset. Cheng *et al*., likely due to their Taiwan CDC affiliation, had access to more comprehensive data than publicly available, as evidenced by their Fig 1 which illustrates detailed daily exposure times for both secondary cases and close contacts. Without access to such comprehensive data, we implemented a weighting function for healthcare contacts occurring after day 8 to more stably capture this pattern. During development, we observed similar healthcare contact patterns emerging naturally without weighting, but these occurrences were inconsistent, leading us to omit these unstable results from our final analysis. The uptick in secondary infection predicted by CovSyn is not present in Cheng *et al*., indicating that CovSyn does not fully reflect the highly successful isolation strategies employed in Taiwan.

It’s important to note that CovSyn functions as a data synthesizer rather than a forecasting model. In our Validation of population-level data section, we focused on evaluating CovSyn’s ability to capture patterns in the observed Taiwan COVID-19 dataset instead of forecasting future disease dynamics. To assess the local transmission, the daily local confirmed cases and daily local deaths are used. Although the Taiwan CDC also provides daily reports on suspected, excluded, and recovered cases, these figures include both local and abroad cases. To maintain consistency in comparison, we exclude these data from our analysis. A key challenge in this simulation is the lack of real infection dates, as such information requires detailed contact tracing, which is not openly reported.

CovSyn achieved high accuracy for cumulative confirmed cases (*R*^2^ = 0.9) across daily, 7-day centered moving average (MA), and 31-day MA, but showed weaker performance for daily data and death-related metrics as shown in Table 3. The inferior performance for death-related metrics stems from the low number of deaths potentially aided by our temporal alignment methodology. We optimized the time shift parameter solely based on cumulative confirmed cases. Daily data metrics performed worse compared to cumulative metrics, as the observed daily counts are low with numerous zeros interspersed with occasional case spikes, reflecting the stochasticity of infection. In contrast, our averaged outcomes from 1,000 Monte Carlo runs are smooth and capture overall trends rather than the random day-to-day fluctuations.

Despite these limitations, Fig 7 demonstrates that the model successfully captured the overall epidemic trajectory. The synthetic local confirmed cases generated by CovSyn exhibited an earlier increase than observed in the Taiwan COVID-19 dataset but closely mirrored the growth phase and ultimately converged slightly below the actual Taiwanese figures. This difference in early trajectory may be attributed to Taiwan’s prompt public health interventions between January and February 2020, including the classification of SARS-CoV-2 as a notifiable disease (January 15) and implementation of the mask rationing system (February 6), which collectively reduced social contacts during this critical period. Regarding local deaths, the CovSyn model produced slightly higher estimates than those recorded in Taiwan. Most observations fell within the 95% confidence intervals except during the initial pre-growth phase (2020-01-16 to 2020-02-16).

Our CovSyn model synthesizes both detailed individual health trajectories and infection spread through contact networks. Several approaches exist for generating other general synthetic healthcare data. Electronic health record (EHR) synthesis models like Synthea [75] excel at generating detailed individual-level data spanning patients’ lifetimes, including comprehensive clinical information such as diagnoses, medications, procedures, and care plans with appropriate temporal progression. More recently, Nikolentzos *et al*. introduced a graph autoencoder model (VGAE) for generating synthetic EHRs [76]. Their model represents each patient’s medical history as a graph structure, where nodes capture patient information, healthcare encounters, conditions, prescriptions, and test results. However, a key limitation of these EHR-focused approaches is their inability to model infection transmission dynamics between patients through contact networks.

In contrast, agent-based models like Covasim, OpenABM-Covid19, and EpiHiper [12, 53, 56] simulate disease spread by incorporating contact networks and transmission probabilities between individuals in different social settings. These models can reproduce daily case trends while maintaining flexibility to analyze COVID-19 transmission under various scenarios. However, they lack training on detailed individual-level clinical data as shown in Table 1 and thus health trajectories, which CovSyn incorporates.

CovSyn can also be utilized to evaluate methods for estimating epidemiological statistics. For instance, we have utilized it to investigate the impact of detection rates on estimating *R*_0_, as well as to compare different estimation methods [77]. This study includes a comparison of *R*_0_ methods that either use only daily confirmed cases or use contact networks to estimate the *R*_0_.

Our model has several limitations. The calibrated data is collected from the period when the original virus was dominant in Taiwan. Therefore, our model does not account for viral variants. Our analysis focuses exclusively on Taiwan, treating it as a closed system isolated from global interactions. Starting February 6, 2020, Taiwanese nationals with travel history to China, Hong Kong, or Macau were required to undergo 14 days of home quarantine, and later on March 19, 2020, Taiwan introduced mandatory monitored quarantine upon arrival for all travellers, so the island was largely isolated. However, the source cases for each of the four local outbreaks came from abroad [78]. Of 661 abroad cases reported during the monitored quarantine were enforeced and before the second outbreak (2020-03-19 to 2021-01-11). Only 10 have been shown to have caused local infections. We do not currently incorporate international mobility data or consider disease transmission between countries, which means we do not consider abroad cases in our simulation. Instead, our focus is on simulating the local epidemic spread. In future work, we plan to extend our model by simulating multiple interconnected countries using separate CovSyn instances, with cross-border transmission rates derived from mobility data to capture international disease spread.

Moreover, our algorithm is restricted to the available data types. CovSyn generates only the degree for each infected individual in the contact network, indicating how many connections each infected individual has (whether to other infected or uninfected individuals). It does not generate information about connections between uninfected individuals, thus providing only a partial view of the overall contact network structure.

We plan to address this limitation in the future by mapping the generated contact network to demographic data and optimizing it accordingly. We may also incorporate individual behavioural changes to reflect agents’ reactions to interventions. Future development of CovSyn could enhance the model by incorporating more comprehensive clinical data, either from private EHRs or from the synthetic healthcare data models mentioned above. Furthermore, population-level data were not incorporated into our optimization framework, *i*.*e*., the residuals between observed daily confirmed cases and daily deaths versus simulated outcomes were not included in the loss function during CovSyn training. This is due to the unavailability of actual infection dates in the Taiwan COVID-19 data. Consequently, we implemented a simple day-shift approach utilizing the bisection method to optimize temporal alignment post-training. Future work will cover integrating population-level data directly into the optimization procedure to enhance simulation fidelity and predictive accuracy. Additionally, due to data limitations, our current model does not distinguish between asymptomatic and symptomatic cases in the course of disease synthesis. In future iterations, we plan to implement separatration between asymptomatic and symptomatic cases, which would enhance the model’s ability to capture the full spectrum of COVID-19 manifestations.

To our knowledge, this work represents one of the early attempts in the epidemiological modelling field to develop a synthetic dataset specifically designed for disease model development, which is calibrated on individual-level data. These ideas have been validated in the systems biology field (*e*.*g*. synthetic gene expression datasets for inference of gene regulatory networks) and published in top journals [23, 79]. The insufficiency of COVID-19 public data has hampered the development of reliable epidemiological forecasting modelling. We synthesized the detailed individual-level data of COVID-19 subjects based on Taiwan public data and research statistics. The synthetic dataset is consistent with the current understanding of the original SARS-CoV-2 and human behavior, and it provides detailed information, such as the specific infection history and contact network, that is not available in public data. We validated the synthetic dataset by comparison to the Taiwan COVID-19 dataset and clinic observations. CovSyn can help generate a detailed, individual-level synthetic COVID-19 dataset for benchmarking modelling and analysis a synthetic pandemic.

## Materials and methods

### Course of disease of COVID-19 subject

A subject’s course can be summarized as susceptible (at risk of infection, S) to infected (infection by SARS-CoV-2 virus, I), infected to recovered (two consecutive negative PCR tests, R) or to death (D). The state I can be further expanded as asymptomatic (infection with no symptoms, I^A^), symptomatic (infection with symptoms, I^S^), and critically ill (ICU admission, I^C^) as shown in Fig 5. Infection can be confirmed by an antigen test or PCR test. The immune status can be determined by an antibody test. The outside inputs such as nonpharmaceutical interventions (NPI) and intensive care units (Care) can decrease the transmission probability from susceptible to infection and from susceptible to death. Vaccination can increase the transition probability from susceptible to immune.

During the infectious period, an individual is likely to cause secondary infections among other susceptible individuals defined as effective contacts. This branching process is illustrated in Fig 8. Person B and C are the effective contacts of person A; person D is infected by person B; and person E to G are infected by person C.

**Fig 8.**
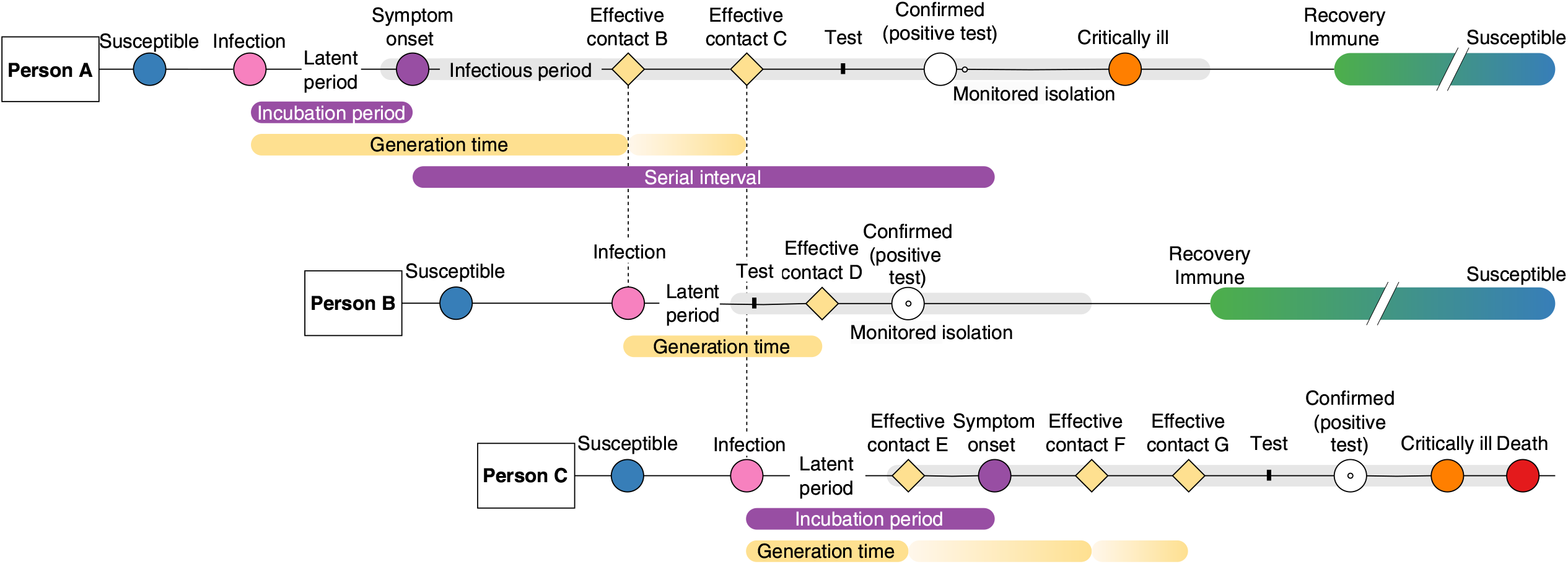
Illustration of the simulated course of disease branching process. Case B and C are the effective contacts of case A, case D get infected by case B, and case E to G are infected by case C.

**Fig 9.**
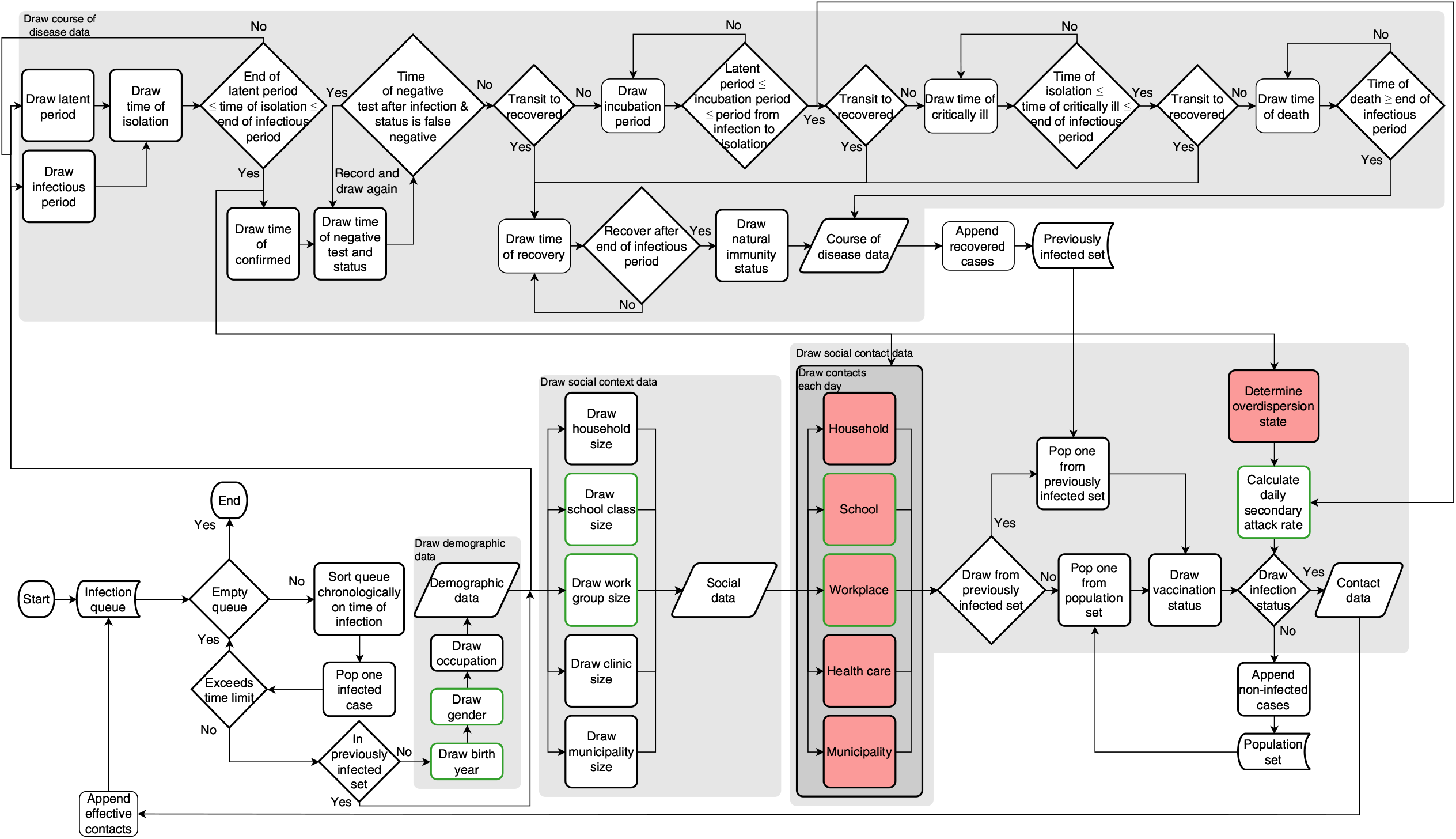
Flow chart of the data synthesis process. The process begins with an infection queue specifying the case IDs and infection time and ends when the queue is empty or the time limit has been reached. The final results are demographic data, social data, contact data, and course of disease data. The boxes with green borders indicate the data related to age. Parameters with no prior knowledge are indicated by red boxes.

The relation of the crucial epidemiological parameters are described in Fig 8. The latent period is defined as the time from infection to the beginning of the infectious period and the incubation period is defined as the time from infection to symptom onset. Each subject has different infectious periods (the grey shaded area in Fig 8), typically starting a few days before the onset of symptom and ending when the viral load is low. The generation time is defined as the time between infection of the infector-infectee pair and the serial interval as the symptom onset interval between the infector-infectee pair. If the case is confirmed and moved to monitored isolation before the end of the infectious period, then the infected person cannot infect others, reducing the spread. Some cases may develop severe symptoms or even die, while most cases recover from the disease and become immune. Eventually, they lose their immunity and return to being susceptible.

### Input data

#### Demographic data

We collected age and gender data in 2022 from the National Development Council (https://pop-proj.ndc.gov.tw/main_en/dataSearch.aspx?uid=78&pid=78). We classified the social data into five different layers: household, school class, workplace, hospital, and municipality. The household/family size and population size in 2021 for each county/city were obtained from the Department of Household Registration, Ministry of Interior (https://www.ris.gov.tw/app/portal/346). The class size for each grade from each county/city was gathered from elementary school to university from the Department of Statistics, Ministry of Education in 2021 (https://depart.moe.edu.tw/ED4500/News_Content.aspx?n=5A930C32CC6C3818&sms=91B3AAE8C6388B96&s=8AF80DB14ADF7370). For the workplace-related data, we collected the employment rate vs age data in 2021 and job category vs industry data in 2020 from the Gender Equality Committee of the Executive Yuan (https://gec.ey.gov.tw/en/) and National Statistics (https://www.stat.gov.tw/ct.asp?xItem=46590&ctNode=3579&mp=4). From the Ministry of Health and Welfare (https://dep.mohw.gov.tw/DOS/cp-5301-62356-113.html), we collected the statistics on the daily medical service volume of hospitals in 2020.

The demographic parameters consist of 25,505 fixed values, which are detailed in an accompanying Excel table named `demographic_parameters.xlsx`. These parameters include population sizes for each city (no. parameters, 20) and probabilities associated with age (101), gender (202), student status by age and gender (202), employment status by age and gender (202), part-time job (32), full-time job (32), family size distribution by city (172), class size distribution by age (2272), workplace size distribution (22094), hospital distribution (14), and hospital size distribution (162).

### Epidemiological parameters

The course of disease parameters, totalling 161, is optimized by the Firefly algorithm. The initial setting, the source of the parameters, the searching boundary, and the final estimation can be found in an Excel table, named `pe_course_of_disease_parameters.xlsx`. Parameters sourced from prior research (142) cover various aspects such as latent period, infectious period, incubation period, time from infection to death, daily secondary attack rates, secondary attack rates within households, schools, workplaces, and healthcare settings, natural immunity rates, vaccine efficacy, and age-specific risk ratios. Parameters derived from the Taiwan structured data (19) pertain to statistics derived from fitted Gamma distributions concerning state transition days and state transition probabilities.

The social contact parameters consist of 37 parameters optimized via the Firefly algorithm. These parameters include empirical settings for contact probabilities across five layers, totalling 35 parameters. Additionally, this set encompasses statistics related to the overdispersion rate. Detailed values of the boundary and optimized values for the course of disease parameters and social contact parameters are available in the Excel table `pe_social_contact_parameters.xlsx`. The top 10% of the fireflies are also saved in the Excel table.

### Individual course of disease and contact tracing data

We have collected the Taiwan COVID-19 dataset containing course of disease data from 579 confirmed cases and their contact tracing information from Taiwanese COVID-19 open datasets. To our knowledge, this dataset represents the most comprehensive compilation of individual-level course of disease data and contact tracing information available publically to date, attributed to the diligent efforts of the Taiwan CDC in disease control and daily data reporting. The details of the data can be found in our article [21].

#### Transmission dynamics data

Transmission dynamics data from Cheng *et al*. [26] were collected. This data contains the number of secondary infected cases and the number of secondary close contacts for 100 source cases with respect to days from onset to first exposure. They categorized the contacts into household, healthcare, and others.

### Preprocessing

#### Initialization dataset

For the disease state transition data, we fitted Gamma distributions to model the duration of state transitions in our dataset. To estimate the confidence intervals for the Gamma distribution parameters, we employed a bootstrap method. These confidence intervals served as upper and lower bounds in our optimization process, with the median of the fitted parameters used as initial values. For transitions with limited data points, such as asymptomatic to recovered, symptomatic to recovered, and critically ill to recovered, we applied standard Gamma distribution fitting techniques.

The analysis focused solely on Taiwan’s main island, excluding its offshore territories. Data on household family sizes were categorized into six groups: 1, 2, 3, 4, 5, and *>* 6. To address the unspecified *>* 6 group, total population data for each city was utilized. The approach assumed a monotonic decrease in household frequency for family sizes ranging from 1 to 12. The distribution of family sizes from 6 to 12 was estimated using constrained least squares optimization with non-negative bounds. This optimization minimized the difference between the observed and predicted values for total population and total family sizes in the *>* 6 group for each city. The method achieved high accuracy, with the worst root mean square error (RMSE) across all cities of 8.2 *×* 10^*−*11^, implying that the cumulative sum of individuals in families agrees for all cities, since Taiwan only has 2.3 *×* 10^7^ people.

The code for processing the demographic data and Taiwan COVID-19 dataset to generate all parameters Excel file is in `parameters_for_initialization.py`. It’s important to note that this code is tailored to our specific dataset and research context. However, similar procedures could be adapted for use with datasets from other specific countries.

#### Training dataset

The infection date in the Taiwan COVID-19 dataset is defined as the first date of contact with the source case. Cases with multiple infection dates are not considered in the analysis. The dataset contains two deaths that lack ICU admission dates. For these cases, we set the time from symptom onset to critically ill status as the time from symptom onset to death, and the time from critically ill status to death to zero.

The informative subset of individuals from Taiwan data contains features such as age, time from infection to symptomatic, time from symptomatic to critically ill, time from symptomatic to critically ill, time from critically ill to recovered, time from critically ill to death, time from the first negative test to confirmed, time from symptomatic to confirmed, and number of effective contacts. We dropped the data of time from infection to recovery since there were only two subjects containing such data. The asymptomatic date is not considered a test feature since it is an oversimplified estimated value and could cause the distortion of the real distribution. Eventually, we used the most informative cases (containing the values of at least 4 features) from the Taiwan COVID-19 dataset and constructed a 121 *×* 9 test data matrix.

Because we are doing multiobjective optimization, we consider not just the energy distance, but also the contact tracing data including the number of secondary infected cases and the number of close contacts for household, healthcare, and others. We set the 68% confidence interval (one standard deviation) for the state transition parameters as the upper and lower bounds for the optimization instead of the 95% confidence interval (two standard deviations) due to the limited data available. Using too wide a confidence interval could lead to optimized parameters deviating significantly from the original disease state transition data. To balance the need for optimization exploration with the desire to remain close to the original distribution, we chose a one-standard deviation range as the confidence interval. This approach allows for parameter adjustment while preventing excessive deviation from the observed data.

### CovSyn—Agent-based model for data synthesis

Taiwan’s public individual dataset contained information about the course of disease and contact tracing for some cases. Most individuals, however, only included part of the information, making statistical analysis and modelling difficult. The insufficiently detailed progression of the disease for each individual motivated us to synthesized individual data based on the collected data and statistics of COVID-19 epidemiological parameters.

Our data synthesis algorithm consists of four main modules: draw demographic data, draw social context data, draw course of disease data, and draw contact data. The flow chart is in Fig 9 and the algorithm is in Algorithm 1. An outbreak is simulated by inputting an individual in the infection queue. Each individual in the infection queue is assigned a case ID, infection time, and previously infected status. As long as the queue is not empty and the outbreak time does not exceed the time limit, the process will keep synthesizing source cases’ detailed information. After sorting the queue based on the infection time, one infected case is poped out from the infection queue tostart synthesizing the demographic data.

#### Algorithm 1

CovSyn

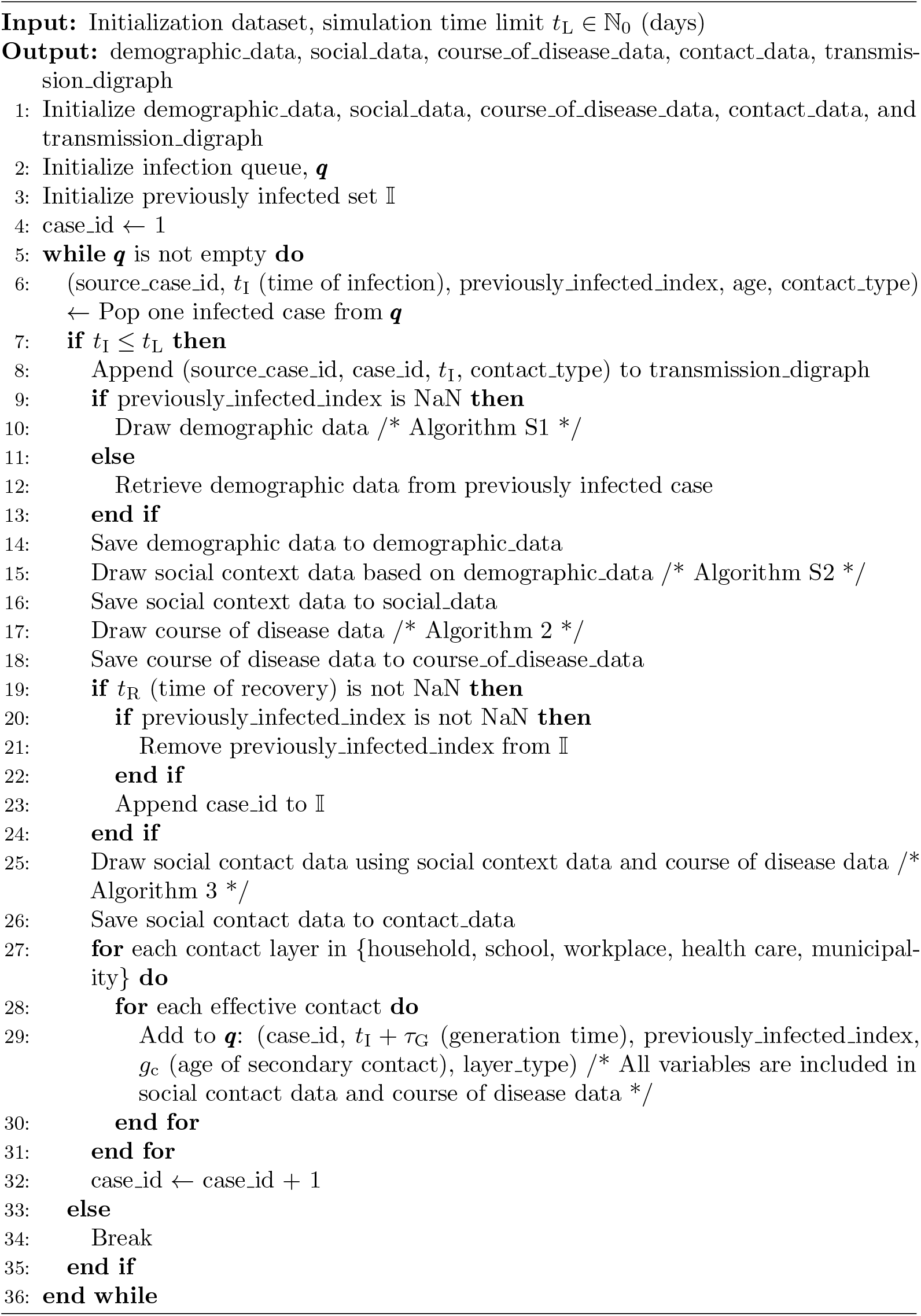

#### Draw demographic and social context data

Demographic data included a person’s age, gender, and occupation. The draw demographic data pseudocode is shown in Algorithm S1. Demographic information will affect the proportion of contacts within different layers, for example, a 50-year-old female is less likely to have school contacts than a 9-year-old girl.

The social context data are categorized into five different layers, which are household, school, workplace, health care, and municipality. The pseudocode is shown in Algorithm S2.

#### Draw course of disease data

The latent period, infectious period, and isolation time were simulated. We set the isolation date to be less than the sum of the latent period and infectious period, since it does not help to isolate the subject cease to be infectious.

Both positive and negative test dates were drawn. The date of the positive test was computed by adding a day shift upon monitored isolation date following a normal distribution with 0 mean and variance 0.3. This is to add randomness so that the positive test date happens a few days before and after the monitored isolation date. The false rate was set to be within 1% to 30% [81]. All the false negatives and true negatives before the infection were saved in the course of disease data.

Following the state transition network shown in Fig 5, an infected person can progress through various disease states. Initially, the person can either become symptomatic or recover directly from an asymptomatic infection. If symptomatic, they may either progress to a critically ill state or recover. Those who become critically ill can either recover or die.

For each transition, we draw a progression time and verify if it fulfils the time constraints. We also assign natural immunity, which affects the secondary attack rate. The complete course of disease for each infected individual is recorded in the infected matrix, allowing us to access all dates of disease progression for any infected person. Our algorithm for simulating the course of disease is outlined in Algorithms 2. Rather than simulating transitions for each disease state on a daily basis, we directly simulate the transition times between states to reduce the computation needed.

##### Algorithm 2

Draw course of disease data

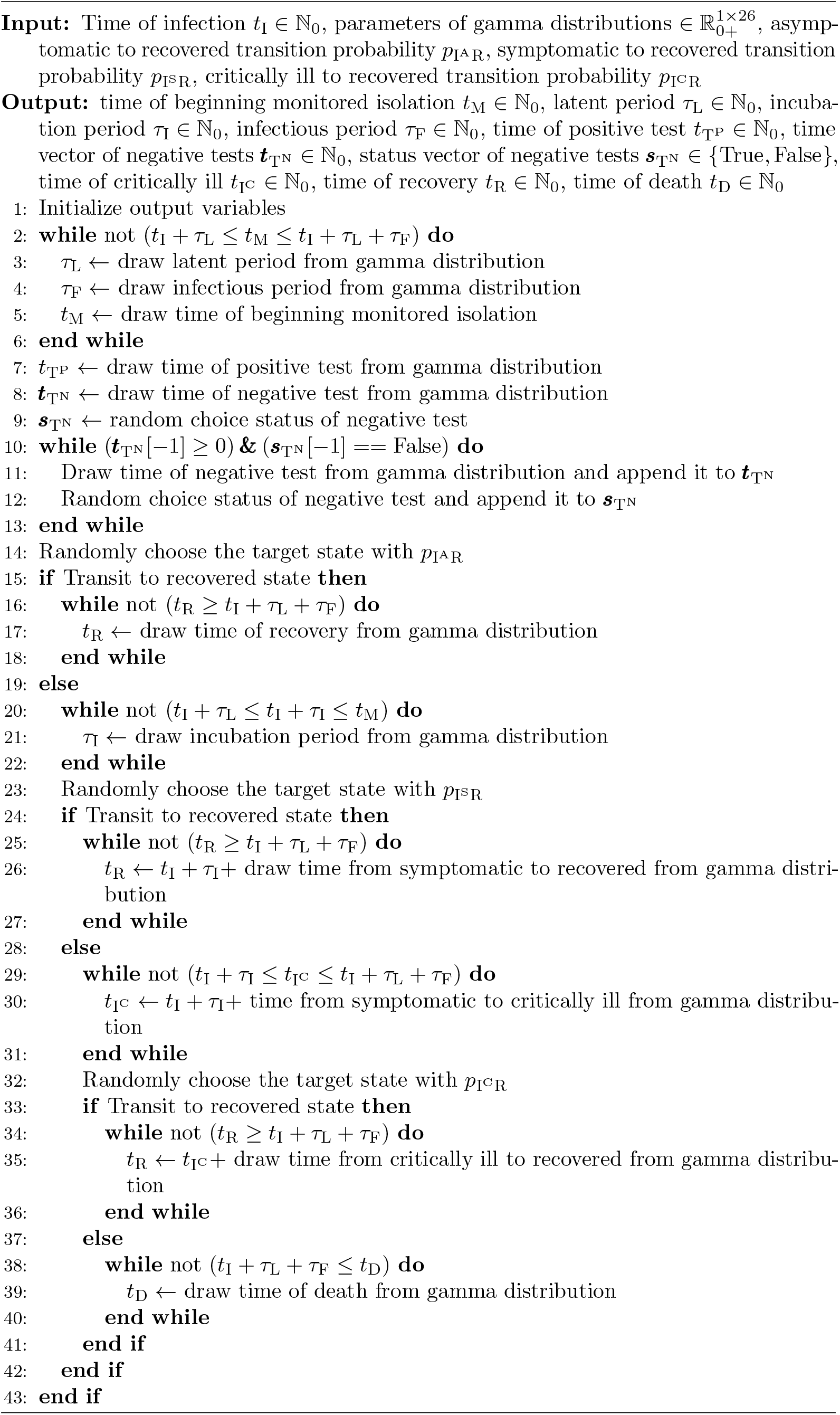

#### Draw social contact data

The daily contacts for each social layer were synthesized using input parameters including social group size, course of disease (specifically incubation period, infectious period, and isolation period), and contact probability. A source case can only have valid contacts during the period from infection to isolation, with each day having a specific contact probability. We assumed that human behaviour regarding close contacts would change in the days following symptom onset, following an exponential trend. To model this, we defined the contact probability function *f*_*p*_*L* for cases with no contact on the previous day by combining two logistic functions. This approach captures the changing likelihood of contact as the disease progresses.

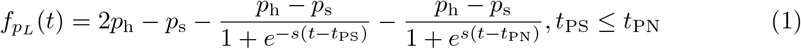

where *p*_h_ and *p*_s_ are the contact probability when healthy and symptomatic. *s* is the steepness, the higher value, the faster the exponential changes. *t*_PS_ and *t*_PN_ represent the temporal phases relative to symptom onset time *t*_I_S with *t*_PS_ marking the beginning of symptom manifestation and *t*_PN_ indicating the transition point where contact patterns begin returning to normal.

For most of the layers, the contact probability decreased to a small number to reflect people avoiding meeting other people after symptom onset and returning to normal contact probability when they felt better. For the healthcare layer, the situation was the opposite, people tended to meet more healthcare workers a few days after symptom onset. The Draw contacts each day code is shown in Algorithm S9. Our exponential trending contact probability function can mimic both the assumption of the change in human behaviour at the beginning of the pandemic and that people learn how to protect themselves and their families by isolating themselves right after the symptom onset.

After the draw contacts each day process, the contact date for each contact in each layer can be simulated. The source case might have multiple contacts either from the previously infected set or from the population set. The vaccination status will be drawn for each contact. All the contact types, vaccination status, natural immunity, and overdispersion state will affect the calculation of the daily secondary attack rate. The natural immunity rate was set to be 0.91 [82]. With the secondary attack rate and contact dates, we can decide which date the contact gets infected, *i*.*e*. the effective contact date. The effective contact list was then sorted by infection time and appended back to the infection queue at the start of the next iteration. Our algorithm for drawing contact data is Algorithm 3.

##### Algorithm 3

Draw social contact data

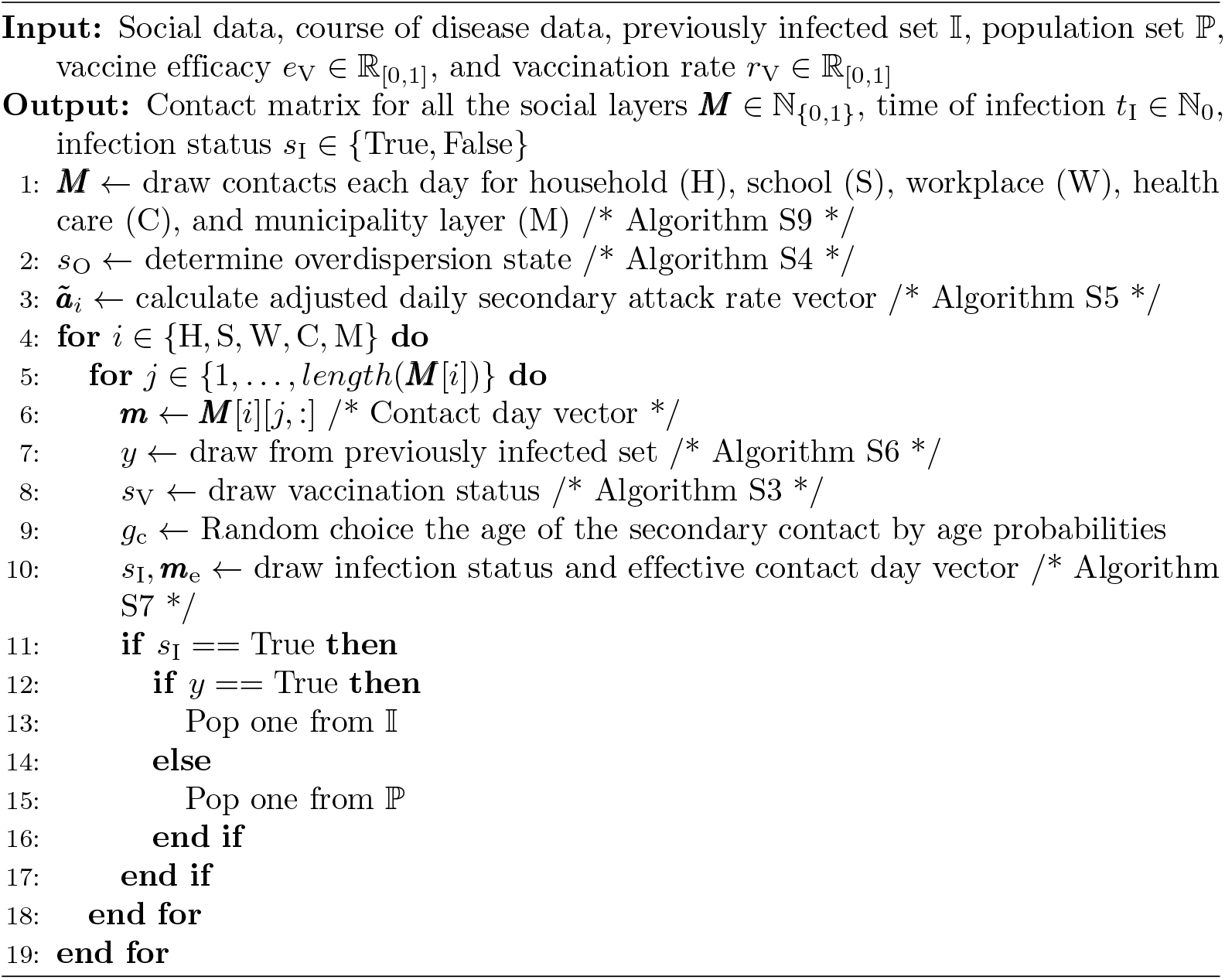

### Firefly optimizer

#### Energy statistics

We apply energy distance statistics to quantify how different our synthetic dataset is from the observed Taiwan COVID-19 dataset. The null hypothesis is that both multidimensional data come from the same distribution. The idea is to make the synthetic dataset similar to the observed Taiwan COVID-19 dataset.

Consider the two datasets 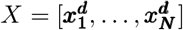 and 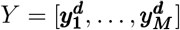, where *N* and *M* are the number of samples. Each of ***x***^***d***^ and ***y***^***d***^ is a *d* dimentional vector. The energy distance is defined as

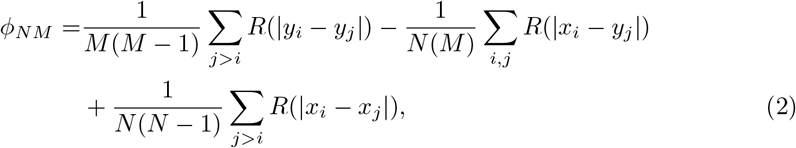

where *R*(*·*) is the distance function. We use the logarithmic distance function

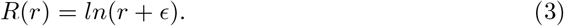

The small positive constant *ϵ* is set to be 10^*−*16^. Since our observed data is a sparse matrix, *i*.*e*. containing many NaN values, we remove the NaN and calculate the Euclidean distances. The missing values are ignored in order to not distort the distribution.

#### Optimization by Firefly algorithm

Due to the complexity of our model, which involves unknown parameters, non-linear behaviour, and multiple local minima, traditional gradient-based methods prove insufficient for exploring the parameter space and achieving accurate estimates. To address this challenge, we have employed the Firefly algorithm, a swarm intelligence optimization method. This approach, detailed by Yang [83], enables our model to avoid local minima and converge closer to the global minimum. Compared to other soft computing methods, the Firefly algorithm efficiently handles NP-hard problems with multi-modal functions [84]. Its mechanism strikes a balance between exploration and exploitation, enhancing the learning process during parameter training.

Parameters without lower and upper bounds from clinical research were perturbed by 30%. The algorithm first assigned fireflies with random positions. Then in each iteration *k*, the current firefly *i* compares with other fireflies and updates its position based on their intensities. If the other firefly *j* is brighter (lower cost), the current firefly position *x*_*i*_ would be altered with the function,

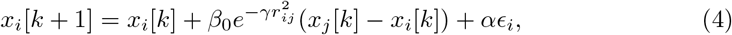

where *r*_*ij*_ is the Euclidean distance between the two fireflies. We set the attractiveness constant *β*_0_ = 1 and light absorption coefficient *γ* = 0.131 to make sure one firefly can ‘see’ 10% to 20% of the fireflies. The random walk parameter, *α*, was set to be 0.97^*k*^ indicating a decreasing value in each iteration. The min-max normalization was implemented to avoid weighting the 198 parameters. One generation is completed when all fireflies update their positions.

The residual sum of squares (RSS) of the synthesis secondary contact number and infection number each day compared to Cheng *et al*.’s observation was calculated [26]. We normalized Cheng’s observation so that the maximum value is 1 and we applied the same normalization to the synthetic secondary contacts. The contact number each day is grouped as *<* 0, 0 *−* 3, 4 *−* 5, 6 *−* 7, 8 *−* 9, and *>* 9 days from onset to the first exposure. In addition, we calculated the RSS of secondary attack rates (*RSS*^*a*^). We applied the same normalization techniques to the secondary attack rates. Based on Cheng *et al*.’s prediction, secondary attack rates in each contact day group have different uncertainty. The reported secondary attack rate with a lower uncertainty range should have a higher weight in the optimization. We thus multiply weights that are set to be the inverse of the uncertainty range to the reported secondary attack rates. The min-max normalization was applied to each layer of the attack weight. The optimization is conducted layer by layer. For the ‘health care’ layer, we observed in the Cheng et al. that the daily contact number increases after day 8 from symptom onset. To capture this phenomenon, we applied a weight of 2 to the daily contacts occurring after day 8.

The energy distance (*E*) is also considered in the optimization to minimize the distance between the Taiwan COVID-19 dataset and our synthetic courses of disease. Finally, the cost function is defined as,

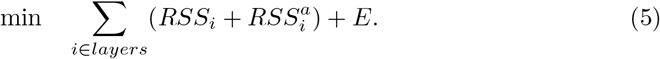

A Linux server with Intel Core i5-7600 CPU, 40G RAM, and Ubuntu 22.04 was used to optimize the 198 parameters and generate the synthetic dataset. The optimization process took approximately 5.1 days for 1,103,479 evaluations with the setting: firefly size 100 and maximum generation 200. The main data synthesis algorithm took 0.4 seconds to generate a dataset with 100 cases.

### Monte Carlo simulations

In this section we detailed the three Monte Carlo simulations that we conducted to evaluate the performance of our model.

#### Course of disease simulation

To study the course of disease statistics shown in Fig 5, we simplified our approach by focusing only on course of disease without saving all the contact networks. We conducted 10,000 simulations, each initialized with a single source case with infection on day 0, and tracked only their course of disease history.

#### Secondary close contact simulation

In this scenario, we designated 100 individuals as source cases as in the experiment setting in Cheng *et al*. [26], All 100 source individuals were initialized with infection on day 0. We tracked both the secondary close contacts and the course of disease of all 100 source cases. The model did not simulate tertiary transmission (contacts of contacts), *i*.*e*., the branching process was not implemented in this case.

#### First outbreak in Taiwan

To compare with Taiwanese population-level data, we performed 1,000 Monte Carlo simulations with a time horizon of 365 days. Each simulation was initialized with the 28 seed infections, which were identified by analyzing infection chains from available contact tracing data in the Taiwan COVID-19 dataset. We selected individuals with the earliest confirmed cases as these source cases. For the epidemiological parameter distributions shown in Fig 3, we utilized the first 100 simulations to generate a total of 3,176 infected individuals.

For the Validation of population-level data section, we compared the full 1,000 simulations with the processed Taiwanese population-level data as the ground truth for this comparison. Our previous work [78] details the preprocessing methodology applied to this Taiwanese population-level data.

A critical aspect of this analysis involved optimizing the temporal alignment between synthetic and observed data through a shift parameter of *x* days. The maximum allowable shift was set to 57 days—the period between the first local confirmed case in Taiwan and December 1, 2020 (the symptom onset date of the first identified case in Wuhan, China [85]). To determine the optimal shift value, we applied the bisection method, iteratively adjusting the parameter *x* and calculating the mean square error (MSE) between the mean of the 1,000 simulated local confirmed cases and the Taiwanese local confirmed cases. Note that this optimization process takes place after the CovSyn model has already been trained, *i*.*e*. this bisection optimization is completely separate from the training process. The shift value that minimized MSE was selected as optimal. This optimal shift value was then applied to all simulated data, including infection day, confirmation day, and death day data.

The validation covered the period from December 31, 2019, to June 16, 2020, examining model performance at daily, weekly, and monthly aggregation levels for both confirmed cases and deaths.

## Data Availability

The CovSyn model, data, and documentation are fully available via GitHub https://github.com/nordlinglab/COVID19-CovSyn

## Acknowledgments

As non-native English speakers, we acknowledge the work of OpenAI, L.L.C. in creating GPT4 and Anthropic in creating Claude 3.7, which helped us improve the readability and language of this article. We would like to thank the Ministry of Science and Technology in Taiwan for their financial support (Grants Number MOST 105-2218-E-006-016-MY2, 105-2911-I-006-518, 107-2634-F-006-009, 110-2222-E-006-010, and National Science and Technology Council 111-2221-E-006-186, 112-2314-B-006-079, and 113-2314-B-006-069). This research was supported in part by Higher Education Suprout Project, Ministry of Education to Headquarters of University Advancement at National Cheng Kung University (NCKU).

