## Supplementary for "CovSyn: an agent-based model for synthesizing COVID-19 course of disease and contact tracing data"

* torbjörn.nordlingnordlinglab.org

**Table S1.** Notation list of the mathematical symbols used in this article. Italic small letters denote variables, bold small letters denote arrays, and Roman font indicates physical quantities.

| Notation | Description | Notation | Description |
|---|---|---|---|
| $t_{\mathrm{L}}$ | Simulation time limit (day) | $\tilde{\boldsymbol{a}}$ | Adjusted daily secondary attack rate vector |
| $t_{\mathrm{S}}$ | Time of becoming susceptible (day) | $\tilde{\boldsymbol{a}}^{\mathrm{a}}$ | Age adjusted daily secondary attack rate vector |
| $t_{\mathrm{I}}$ | Time of infection (day) | $\boldsymbol{m}$ | Contact day vector |
| $t_{\mathrm{I^S}}$ | Time of symptom onset (day) | $c$ | Contacts number |
| $t_{\mathrm{I^C}}$ | Time of critically ill (day) | $n_d$ | Number of days |
| $t_{\mathrm{D}}$ | Time of death (day) | $\boldsymbol{m}_{\mathrm{e}}$ | Effective contact day vector |
| $t_{\mathrm{R}}$ | Time of recovery (day) | $s_{\mathrm{I}}$ | Infection status |
| $t_{\mathrm{M}}$ | Time of beginning monitored isolation (day) | $g_{\mathrm{c}}$ | Age of the secondary contact |
| $t_{\mathrm{T}}$ | Time of COVID-19 test (day) | $\boldsymbol{q}$ | Infection queue |
| $t_{\mathrm{T^P}}$ | Time confirmed/positive COVID-19 test (day) | $\mathbb{I}$ | Previously infected set |
| $\boldsymbol{t}_{\mathrm{T^N}}$ | Time vector of negative COVID-19 tests (day) | $\mathbb{P}$ | Population set |
| $\boldsymbol{s}_{\mathrm{T^N}}$ | Status vector of negative COVID-19 tests (boolean) | $\boldsymbol{d}_{\mathrm{C}}$ | Daily confirmed cases |
| $\tau_{\mathrm{I}}$ | Incubation period (days) | $\boldsymbol{d}_{\mathrm{D}}$ | Daily deaths |
| $\tau_{\mathrm{F}}$ | Infectious period (days) | $\boldsymbol{d}_{\mathrm{M}}$ | Daily hospitalizations (monitored isolation) |
| $\tau_{\mathrm{M}}$ | Period from infection to monitored isolation (days) | $f_{p_L}$ | Contact probability function (Eq. 1) |
| $\tau_{\mathrm{G}}$ | Generation time (days) | $\boldsymbol{p}$ | Daily contact probability vector |
| $\tau_{\mathrm{S}}$ | Serial interval (days) | $\boldsymbol{r}_a$ | Age-related risk ratio vector of the secondary attack rate |
| $\tau_{\mathrm{L}}$ | Latent period (days) | $s_{\mathrm{N}}$ | Natural immunity status (boolean) |
| $n_{\mathrm{H}}$ | Household size (people) | $s_{\mathrm{V}}$ | Vaccination status (boolean) |
| $n_{\mathrm{S}}$ | School class size (people) | $p$ | Probability of contact |
| $n_{\mathrm{W}}$ | Work group size (people) | $p_{\mathrm{c}}$ | Consecutive daily contact probability |
| $n_{\mathrm{C}}$ | Clinic size (people) | $p_{\mathrm{h}}$ | Contact probability when healthy |
| $n_{\mathrm{M}}$ | Municipality size (people) | $p_{\mathrm{s}}$ | Contact probability when symptomatic |
| $n$ | Sum of social context size (people), $n_{\mathrm{H}} + n_{\mathrm{S}} + n_{\mathrm{W}} + n_{\mathrm{C}} + n_{\mathrm{M}}$ | $s$ | Steepness of the logistic contact probability function |
| $\boldsymbol{a}$ | Daily secondary attack rate vector | $t_{\mathrm{PS}}$ | Phase relative to symptom onset for symptomatic (days) |
| $\boldsymbol{a}_{\mathrm{H}}$ | $\boldsymbol{a}$ for household layer | $t_{\mathrm{PN}}$ | Phase relative to symptom onset for resuming normal social context (days) |
| $\boldsymbol{a}_{\mathrm{S}}$ | $\boldsymbol{a}$ for school layer | $s_{\mathrm{O}}$ | Overdispersion state |
| $\boldsymbol{a}_{\mathrm{W}}$ | $\boldsymbol{a}$ for workplace layer | $\boldsymbol{M}$ | Contact matrix for all the social layers |
| $\boldsymbol{a}_{\mathrm{C}}$ | $\boldsymbol{a}$ for health care layer | | |
| $\boldsymbol{a}_{\mathrm{M}}$ | $\boldsymbol{a}$ for municipality layer | | |

**Table S2.** Notation list of the mathematical symbols of the contact probability parameters used in this article. Italic small letters denote variables. Roman font indicates physical quantities. Estimate is the value yielded by the best firefly and used in Monte Carlo simulations and analysis, while range is the range of values among the set of best fireflies. The initial is the range of initial guesses in the initial population of 100 fireflies, and the constraints are limits enforced.

| Notation | Description | Estimate | Range | Initial | Constraints |
|---|---|---|---|---|---|
| $p_{\mathrm{H}}$ | $p$ within household layer | 0.98 | (0.98, 0.98) | (0.95, 1.00) | (0.95, 1.00) |
| $p_{\mathrm{cH}}$ | $p_{\mathrm{c}}$ within household layer | 0.40 | (0.40, 0.41) | (0.40, 0.99) | (0.40, 1.00) |
| $p_{\mathrm{hH}}$ | $p_{\mathrm{h}}$ within household layer | 0.06 | (0.05, 0.06) | (0.05, 0.50) | (0.05, 0.50) |
| $p_{\mathrm{sH}}$ | $p_{\mathrm{s}}$ within household layer | 0.02 | (0.02, 0.02) | (0.01, 0.10) | (0.01, 0.10) |
| $s_{\mathrm{H}}$ | $s$ within household layer | 3.76 | (3.76, 3.89) | (1.24, 19.98) | (1.00, 20.00) |
| $t_{\mathrm{PSH}}$ | $t_{\mathrm{PS}}$ within household layer | 4.74 | (4.68, 4.77) | (0.10, 9.90) | (0.00, 10.00) |
| $t_{\mathrm{PNH}}$ | $t_{\mathrm{PN}}$ within household layer | 7.14 | (7.11, 7.17) | (0.07, 9.84) | (0.00, 10.00) |
| $p_{\mathrm{S}}$ | $p$ within school layer | 0.80 | (0.80, 0.81) | (0.11, 0.97) | (0.10, 1.00) |
| $p_{\mathrm{cS}}$ | $p_{\mathrm{c}}$ within school layer | 0.91 | (0.91, 0.91) | (0.50, 0.99) | (0.50, 1.00) |
| $p_{\mathrm{hS}}$ | $p_{\mathrm{h}}$ within school layer | 0.01 | (0.01, 0.02) | (0.01, 0.50) | (0.01, 0.50) |
| $p_{\mathrm{sS}}$ | $p_{\mathrm{s}}$ within school layer | 0.04 | (0.04, 0.04) | (0.00, 0.05) | (0.00, 0.05) |
| $s_{\mathrm{S}}$ | $s$ within school layer | 6.73 | (6.72, 6.78) | (1.06, 9.45) | (1.00, 10.00) |
| $t_{\mathrm{PSS}}$ | $t_{\mathrm{PS}}$ within school layer | 10.00 | (9.93, 10.00) | (0.10, 9.96) | (0.00, 10.00) |
| $t_{\mathrm{PNS}}$ | $t_{\mathrm{PN}}$ within school layer | 1.73 | (1.71, 1.77) | (0.00, 9.90) | (0.00, 10.00) |
| $p_{\mathrm{W}}$ | $p$ within workplace layer | 0.22 | (0.21, 0.22) | (0.10, 1.00) | (0.10, 1.00) |
| $p_{\mathrm{cW}}$ | $p_{\mathrm{c}}$ within workplace layer | 0.56 | (0.56, 0.57) | (0.50, 1.00) | (0.50, 1.00) |
| $p_{\mathrm{hW}}$ | $p_{\mathrm{h}}$ within workplace layer | 0.04 | (0.04, 0.04) | (0.01, 0.49) | (0.01, 0.50) |
| $p_{\mathrm{sW}}$ | $p_{\mathrm{s}}$ within workplace layer | 0.02 | (0.02, 0.02) | (0.00, 0.05) | (0.00, 0.05) |
| $s_{\mathrm{W}}$ | $s$ within workplace layer | 2.76 | (2.76, 2.87) | (1.07, 9.78) | (1.00, 10.00) |
| $t_{\mathrm{PSW}}$ | $t_{\mathrm{PS}}$ within workplace layer | 2.78 | (2.69, 2.79) | (0.00, 10.00) | (0.00, 10.00) |
| $t_{\mathrm{PNW}}$ | $t_{\mathrm{PN}}$ within workplace layer | 6.84 | (6.82, 6.87) | (0.03, 9.84) | (0.00, 10.00) |
| $p_{\mathrm{C}}$ | $p$ within heath care layer | 0.01 | (0.01, 0.01) | (0.01, 0.01) | (0.01, 0.01) |
| $p_{\mathrm{cC}}$ | $p_{\mathrm{c}}$ within heath care layer | 0.85 | (0.84, 0.85) | (0.51, 0.99) | (0.50, 1.00) |
| $p_{\mathrm{hC}}$ | $p_{\mathrm{h}}$ within heath care layer | 0.00 | (0.00, 0.00) | (0.00, 0.01) | (0.00, 0.01) |
| $p_{\mathrm{sC}}$ | $p_{\mathrm{s}}$ within heath care layer | 0.72 | (0.72, 0.72) | (0.60, 1.00) | (0.60, 1.00) |
| $s_{\mathrm{C}}$ | $s$ within heath care layer | 4.23 | (4.17, 4.23) | (1.11, 9.92) | (1.00, 10.00) |
| $t_{\mathrm{PSC}}$ | $t_{\mathrm{PS}}$ within heath care layer | 9.37 | (9.35, 9.39) | (0.02, 9.94) | (0.00, 10.00) |
| $t_{\mathrm{PNC}}$ | $t_{\mathrm{PN}}$ within heath care layer | 4.38 | (4.37, 4.44) | (0.16, 9.93) | (0.00, 10.00) |
| $p_{\mathrm{M}}$ | $p$ within municipality layer | 0.00 | (0.00, 0.00) | (0.00, 0.00) | (0.00, 0.00) |
| $p_{\mathrm{cM}}$ | $p_{\mathrm{c}}$ within municipality layer | 0.57 | (0.57, 0.57) | (0.51, 1.00) | (0.50, 1.00) |
| $p_{\mathrm{hM}}$ | $p_{\mathrm{h}}$ within municipality layer | 0.01 | (0.01, 0.01) | (0.01, 0.10) | (0.01, 0.10) |
| $p_{\mathrm{sM}}$ | $p_{\mathrm{s}}$ within municipality layer | 0.05 | (0.05, 0.05) | (0.00, 0.05) | (0.00, 0.05) |
| $s_{\mathrm{M}}$ | $s$ within municipality layer | 3.37 | (3.33, 3.40) | (1.20, 9.84) | (1.00, 10.00) |
| $t_{\mathrm{PSM}}$ | $t_{\mathrm{PS}}$ within municipality layer | 7.41 | (7.33, 7.44) | (0.12, 9.77) | (0.00, 10.00) |
| $t_{\mathrm{PNM}}$ | $t_{\mathrm{PN}}$ within municipality layer | 9.90 | (9.89, 9.95) | (0.02, 9.92) | (0.00, 10.00) |
| $r_{\mathrm{O}}$ | Overdispersion rate | 0.17 | (0.17, 0.17) | (0.00, 0.19) | (0.00, 0.20) |
| $w_{\mathrm{O}}$ | Overdispersion weight | 6.36 | (6.28, 6.41) | (1.12, 19.36) | (1.00, 20.00) |

**Table S3.** Notation list of the mathematical symbols of the epidemiological parameters. Italic small letters denote variables. Roman font indicates physical quantities. The shape, scale, and location parameters define the gamma distributions. Estimate is the value yielded by the best firefly and used in Monte Carlo simulations and analysis, while range is the range of values among the set of best fireflies. The initial is the range of initial guesses in the initial population of 100 fireflies, and the constraints are limits enforced.

| Notation | Description | Estimate | Range | Initial | Constraints |
|---|---|---|---|---|---|
| $k_{\mathrm{L}}$ | Latent period shape | 3.35 | (3.34, 3.36) | (3.35, 5.11) | (3.32, 5.13) |
| $\theta_{\mathrm{L}}$ | Latent period scale | 1.12 | (1.12, 1.12) | (1.08, 1.67) | (1.06, 1.67) |
| $k_{\mathrm{F}}$ | Infectious period shape | 5.96 | (5.96, 5.98) | (2.01, 5.94) | (2.00, 6.00) |
| $\theta_{\mathrm{F}}$ | Infectious period scale | 3.48 | (3.47, 3.49) | (1.00, 3.50) | (1.00, 3.50) |
| $k_{\mathrm{I}}$ | Incubation period shape | 0.76 | (0.75, 0.76) | (0.74, 2.92) | (0.73, 2.93) |
| $\theta_{\mathrm{I}}$ | Incubation period scale | 1.67 | (1.66, 1.67) | (1.61, 8.73) | (1.60, 8.79) |
| $k_{\mathrm{I^S M}}$ | Symptom onset to monitored isolation shape | 0.81 | (0.80, 0.81) | (0.81, 1.58) | (0.79, 1.58) |
| $\theta_{\mathrm{I^S M}}$ | Symptom onset to monitored isolation scale | 6.62 | (6.62, 6.63) | (4.39, 6.91) | (4.39, 6.92) |
| $\mu_{\mathrm{I^S M}}$ | Symptom onset to monitored isolation location | 1.00 | (1.00, 1.00) | (0.93, 1.00) | (0.93, 1.00) |
| $k_{\mathrm{I^A R}}$ | Asymptomatic to recovered shape | 5.04 | (5.03, 5.04) | (4.62, 6.90) | (4.62, 6.93) |
| $\theta_{\mathrm{I^A R}}$ | Asymptomatic to recovered scale | 8.16 | (8.16, 8.18) | (7.24, 10.78) | (7.20, 10.80) |
| $\mu_{\mathrm{I^A R}}$ | Asymptomatic to recovered location | 0.00 | (0.00, 0.00) | (0.00, 0.00) | (0.00, 0.00) |
| $k_{\mathrm{I^S I^C}}$ | Symptom onset to critically ill shape | 1.15 | (1.15, 1.16) | (0.54, 1.93) | (0.53, 1.94) |
| $\theta_{\mathrm{I^S I^C}}$ | Symptom onset to critically ill scale | 5.38 | (5.36, 5.42) | (4.12, 8.98) | (4.07, 8.98) |
| $\mu_{\mathrm{I^S I^C}}$ | Symptom onset to critically ill location | -0.12 | (-0.12, -0.11) | (-0.61, -0.01) | (-0.61, -0.00) |
| $k_{\mathrm{I^S R}}$ | Symptom onset to recovered shape | 2.47 | (2.47, 2.49) | (1.01, 3.35) | (0.97, 3.35) |
| $\theta_{\mathrm{I^S R}}$ | Symptom onset to recovered scale | 7.56 | (7.50, 7.58) | (7.01, 17.21) | (6.96, 17.22) |
| $\mu_{\mathrm{I^S R}}$ | Symptom onset to recovered location | 3.96 | (3.94, 3.98) | (3.36, 7.97) | (3.35, 8.05) |
| $k_{\mathrm{I^C R}}$ | Critically ill to recovered shape | 1.65 | (1.65, 1.65) | (1.23, 1.83) | (1.22, 1.84) |
| $\theta_{\mathrm{I^C R}}$ | Critically ill to recovered scale | 21.14 | (21.12, 21.18) | (14.16, 21.18) | (14.12, 21.18) |
| $\mu_{\mathrm{I^C R}}$ | Critically ill to recovered location | 9.05 | (9.02, 9.07) | (7.42, 11.03) | (7.36, 11.04) |
| $k_{\mathrm{I^A D}}$ | Asymptomatic to death shape | 2.99 | (2.99, 2.99) | (2.96, 3.16) | (2.96, 3.17) |
| $\theta_{\mathrm{I^A D}}$ | Asymptomatic to death scale | 6.57 | (6.56, 6.57) | (6.26, 6.67) | (6.25, 6.67) |
| $k_{\mathrm{NC}}$ | Negative test to confirmed shape | 1.36 | (1.35, 1.36) | (0.63, 2.41) | (0.60, 2.42) |
| $\theta_{\mathrm{NC}}$ | Negative test to confirmed scale | 14.23 | (14.22, 14.25) | (5.88, 14.33) | (5.84, 14.43) |
| $\mu_{\mathrm{NC}}$ | Negative test to confirmed location | 1.87 | (1.87, 1.88) | (1.22, 3.00) | (1.20, 3.00) |
| $r_a$(0-19) | Age-related risk ratio of the secondary attack rate for age group 0-19 years old | 0.03 | (0.02, 0.03) | (0.02, 0.99) | (0.00, 1.00) |
| $r_a$(20-39) | Age-related risk ratio of the secondary attack rate for age group 20-39 years old | 0.14 | (0.13, 0.14) | (0.00, 1.99) | (0.00, 2.00) |
| $r_a$(40-59) | Age-related risk ratio of the secondary attack rate for age group 40-59 years old | 0.93 | (0.91, 0.95) | (0.78, 6.08) | (0.78, 6.14) |
| $r_a$(60+) | Age-related risk ratio of the secondary attack rate for age group 60+ years old | 0.47 | (0.47, 0.52) | (0.46, 6.96) | (0.44, 6.97) |
| $r_{\mathrm{N}}$ | Natural immunity rate | 0.93 | (0.93, 0.93) | (0.87, 0.93) | (0.87, 0.93) |
| $r_{\mathrm{V}}$ | Vaccination rate | 0.00 | (0.00, 0.00) | (0.00, 0.00) | (0.00, 0.00) |
| $e_{\mathrm{V}}$ | Vaccine efficacy | 0.90 | (0.90, 0.90) | (0.88, 0.92) | (0.88, 0.92) |

**Table S4.** Notation list of the mathematical symbols of the day-specific secondary attack rates $a_{\mathrm{H}}(t)$ in the household layer for day $t$ relative to symptom onset. Italic small letters denote variables. Roman font indicates physical quantities. Estimate is the value yielded by the best firefly and used in Monte Carlo simulations and analysis, while range is the range of values among the set of best fireflies. The initial is the range of initial guesses in the initial population of 100 fireflies, and the constraints are limits enforced.

| Notation | Description | Estimate | Range | Initial | Constraints |
|---|---|---|---|---|---|
| $a_{\mathrm{H}}(-14)$ | Day -14 relative to symptom onset | 0.04 | (0.04, 0.04) | (0.04, 0.19) | (0.03, 0.19) |
| $a_{\mathrm{H}}(-13)$ | Day -13 relative to symptom onset | 0.07 | (0.07, 0.07) | (0.06, 0.15) | (0.06, 0.15) |
| $a_{\mathrm{H}}(-12)$ | Day -12 relative to symptom onset | 0.12 | (0.12, 0.12) | (0.09, 0.13) | (0.09, 0.13) |
| $a_{\mathrm{H}}(-11)$ | Day -11 relative to symptom onset | 0.09 | (0.09, 0.09) | (0.09, 0.14) | (0.09, 0.14) |
| $a_{\mathrm{H}}(-10)$ | Day -10 relative to symptom onset | 0.12 | (0.12, 0.12) | (0.08, 0.16) | (0.08, 0.16) |
| $a_{\mathrm{H}}(-9)$ | Day -9 relative to symptom onset | 0.12 | (0.12, 0.12) | (0.07, 0.15) | (0.07, 0.15) |
| $a_{\mathrm{H}}(-8)$ | Day -8 relative to symptom onset | 0.10 | (0.10, 0.10) | (0.07, 0.12) | (0.07, 0.12) |
| $a_{\mathrm{H}}(-7)$ | Day -7 relative to symptom onset | 0.08 | (0.08, 0.08) | (0.07, 0.10) | (0.07, 0.10) |
| $a_{\mathrm{H}}(-6)$ | Day -6 relative to symptom onset | 0.08 | (0.08, 0.08) | (0.06, 0.09) | (0.06, 0.09) |
| $a_{\mathrm{H}}(-5)$ | Day -5 relative to symptom onset | 0.07 | (0.07, 0.07) | (0.06, 0.09) | (0.06, 0.09) |
| $a_{\mathrm{H}}(-4)$ | Day -4 relative to symptom onset | 0.09 | (0.09, 0.09) | (0.07, 0.10) | (0.07, 0.10) |
| $a_{\mathrm{H}}(-3)$ | Day -3 relative to symptom onset | 0.10 | (0.10, 0.10) | (0.09, 0.11) | (0.09, 0.11) |
| $a_{\mathrm{H}}(-2)$ | Day -2 relative to symptom onset | 0.10 | (0.10, 0.11) | (0.10, 0.13) | (0.10, 0.13) |
| $a_{\mathrm{H}}(-1)$ | Day -1 relative to symptom onset | 0.13 | (0.13, 0.13) | (0.11, 0.15) | (0.11, 0.15) |
| $a_{\mathrm{H}}(0)$ | Day 0 relative to symptom onset | 0.13 | (0.12, 0.13) | (0.11, 0.15) | (0.11, 0.15) |
| $a_{\mathrm{H}}(1)$ | Day 1 relative to symptom onset | 0.12 | (0.12, 0.12) | (0.12, 0.14) | (0.12, 0.15) |
| $a_{\mathrm{H}}(2)$ | Day 2 relative to symptom onset | 0.12 | (0.12, 0.12) | (0.11, 0.14) | (0.11, 0.14) |
| $a_{\mathrm{H}}(3)$ | Day 3 relative to symptom onset | 0.11 | (0.11, 0.11) | (0.10, 0.13) | (0.10, 0.13) |
| $a_{\mathrm{H}}(4)$ | Day 4 relative to symptom onset | 0.10 | (0.10, 0.10) | (0.09, 0.12) | (0.09, 0.12) |
| $a_{\mathrm{H}}(5)$ | Day 5 relative to symptom onset | 0.12 | (0.12, 0.12) | (0.09, 0.12) | (0.09, 0.12) |
| $a_{\mathrm{H}}(6)$ | Day 6 relative to symptom onset | 0.11 | (0.11, 0.11) | (0.08, 0.11) | (0.08, 0.11) |
| $a_{\mathrm{H}}(7)$ | Day 7 relative to symptom onset | 0.10 | (0.10, 0.10) | (0.08, 0.11) | (0.08, 0.11) |
| $a_{\mathrm{H}}(8)$ | Day 8 relative to symptom onset | 0.09 | (0.09, 0.09) | (0.08, 0.11) | (0.08, 0.11) |
| $a_{\mathrm{H}}(9)$ | Day 9 relative to symptom onset | 0.09 | (0.09, 0.09) | (0.07, 0.12) | (0.07, 0.12) |
| $a_{\mathrm{H}}(10)$ | Day 10 relative to symptom onset | 0.08 | (0.08, 0.08) | (0.06, 0.14) | (0.06, 0.14) |

**Table S5.** Notation list of the mathematical symbols of the day-specific secondary attack rates $a_{\mathrm{S}}(t)$ in the school layer for day $t$ relative to symptom onset. Italic small letters denote variables. Roman font indicates physical quantities. Estimate is the value yielded by the best firefly and used in Monte Carlo simulations and analysis, while range is the range of values among the set of best fireflies. The initial is the range of initial guesses in the initial population of 100 fireflies, and the constraints are limits enforced.

| Notation | Description | Estimate | Range | Initial | Constraints |
|---|---|---|---|---|---|
| $a_{\mathrm{S}}(-14)$ | Day -14 relative to symptom onset | 0.05 | (0.05, 0.05) | (0.00, 0.06) | (0.00, 0.06) |
| $a_{\mathrm{S}}(-13)$ | Day -13 relative to symptom onset | 0.04 | (0.04, 0.04) | (0.01, 0.05) | (0.01, 0.05) |
| $a_{\mathrm{S}}(-12)$ | Day -12 relative to symptom onset | 0.02 | (0.02, 0.02) | (0.01, 0.04) | (0.01, 0.04) |
| $a_{\mathrm{S}}(-11)$ | Day -11 relative to symptom onset | 0.02 | (0.02, 0.02) | (0.01, 0.04) | (0.01, 0.05) |
| $a_{\mathrm{S}}(-10)$ | Day -10 relative to symptom onset | 0.02 | (0.02, 0.02) | (0.01, 0.05) | (0.01, 0.05) |
| $a_{\mathrm{S}}(-9)$ | Day -9 relative to symptom onset | 0.03 | (0.03, 0.03) | (0.01, 0.05) | (0.01, 0.05) |
| $a_{\mathrm{S}}(-8)$ | Day -8 relative to symptom onset | 0.01 | (0.01, 0.01) | (0.01, 0.04) | (0.01, 0.04) |
| $a_{\mathrm{S}}(-7)$ | Day -7 relative to symptom onset | 0.03 | (0.03, 0.03) | (0.01, 0.03) | (0.01, 0.03) |
| $a_{\mathrm{S}}(-6)$ | Day -6 relative to symptom onset | 0.02 | (0.02, 0.02) | (0.01, 0.03) | (0.01, 0.03) |
| $a_{\mathrm{S}}(-5)$ | Day -5 relative to symptom onset | 0.02 | (0.02, 0.02) | (0.01, 0.03) | (0.01, 0.03) |
| $a_{\mathrm{S}}(-4)$ | Day -4 relative to symptom onset | 0.02 | (0.02, 0.02) | (0.01, 0.03) | (0.01, 0.03) |
| $a_{\mathrm{S}}(-3)$ | Day -3 relative to symptom onset | 0.02 | (0.02, 0.02) | (0.01, 0.03) | (0.01, 0.03) |
| $a_{\mathrm{S}}(-2)$ | Day -2 relative to symptom onset | 0.04 | (0.04, 0.04) | (0.01, 0.04) | (0.01, 0.04) |
| $a_{\mathrm{S}}(-1)$ | Day -1 relative to symptom onset | 0.02 | (0.02, 0.02) | (0.01, 0.04) | (0.01, 0.05) |
| $a_{\mathrm{S}}(0)$ | Day 0 relative to symptom onset | 0.03 | (0.03, 0.04) | (0.01, 0.05) | (0.01, 0.05) |
| $a_{\mathrm{S}}(1)$ | Day 1 relative to symptom onset | 0.03 | (0.03, 0.03) | (0.01, 0.05) | (0.01, 0.05) |
| $a_{\mathrm{S}}(2)$ | Day 2 relative to symptom onset | 0.03 | (0.03, 0.03) | (0.01, 0.04) | (0.01, 0.04) |
| $a_{\mathrm{S}}(3)$ | Day 3 relative to symptom onset | 0.02 | (0.02, 0.02) | (0.01, 0.04) | (0.01, 0.04) |
| $a_{\mathrm{S}}(4)$ | Day 4 relative to symptom onset | 0.02 | (0.02, 0.02) | (0.01, 0.04) | (0.01, 0.04) |
| $a_{\mathrm{S}}(5)$ | Day 5 relative to symptom onset | 0.03 | (0.03, 0.03) | (0.01, 0.04) | (0.01, 0.04) |
| $a_{\mathrm{S}}(6)$ | Day 6 relative to symptom onset | 0.02 | (0.02, 0.02) | (0.01, 0.03) | (0.01, 0.03) |
| $a_{\mathrm{S}}(7)$ | Day 7 relative to symptom onset | 0.02 | (0.02, 0.02) | (0.01, 0.03) | (0.01, 0.03) |
| $a_{\mathrm{S}}(8)$ | Day 8 relative to symptom onset | 0.03 | (0.03, 0.03) | (0.01, 0.03) | (0.01, 0.03) |
| $a_{\mathrm{S}}(9)$ | Day 9 relative to symptom onset | 0.03 | (0.03, 0.03) | (0.01, 0.04) | (0.01, 0.04) |
| $a_{\mathrm{S}}(10)$ | Day 10 relative to symptom onset | 0.02 | (0.02, 0.02) | (0.01, 0.04) | (0.01, 0.04) |

**Table S6.** Notation list of the mathematical symbols of the day-specific secondary attack rates $a_{\mathrm{W}}(t)$ in the workplace layer for day $t$ relative to symptom onset. Italic small letters denote variables. Roman font indicates physical quantities. Estimate is the value yielded by the best firefly and used in Monte Carlo simulations and analysis, while range is the range of values among the set of best fireflies. The initial is the range of initial guesses in the initial population of 100 fireflies, and the constraints are limits enforced.

| Notation | Description | Estimate | Range | Initial | Constraints |
|---|---|---|---|---|---|
| $a_{\mathrm{W}}(-14)$ | Day -14 relative to symptom onset | 0.14 | (0.14, 0.14) | (0.00, 0.22) | (0.00, 0.22) |
| $a_{\mathrm{W}}(-13)$ | Day -13 relative to symptom onset | 0.15 | (0.15, 0.15) | (0.01, 0.17) | (0.01, 0.17) |
| $a_{\mathrm{W}}(-12)$ | Day -12 relative to symptom onset | 0.11 | (0.11, 0.11) | (0.01, 0.15) | (0.01, 0.15) |
| $a_{\mathrm{W}}(-11)$ | Day -11 relative to symptom onset | 0.15 | (0.15, 0.15) | (0.01, 0.17) | (0.01, 0.17) |
| $a_{\mathrm{W}}(-10)$ | Day -10 relative to symptom onset | 0.09 | (0.09, 0.09) | (0.01, 0.19) | (0.01, 0.19) |
| $a_{\mathrm{W}}(-9)$ | Day -9 relative to symptom onset | 0.08 | (0.08, 0.08) | (0.01, 0.17) | (0.01, 0.18) |
| $a_{\mathrm{W}}(-8)$ | Day -8 relative to symptom onset | 0.09 | (0.09, 0.09) | (0.01, 0.14) | (0.01, 0.14) |
| $a_{\mathrm{W}}(-7)$ | Day -7 relative to symptom onset | 0.10 | (0.10, 0.10) | (0.01, 0.11) | (0.01, 0.11) |
| $a_{\mathrm{W}}(-6)$ | Day -6 relative to symptom onset | 0.05 | (0.04, 0.05) | (0.01, 0.10) | (0.01, 0.11) |
| $a_{\mathrm{W}}(-5)$ | Day -5 relative to symptom onset | 0.01 | (0.01, 0.01) | (0.01, 0.11) | (0.01, 0.11) |
| $a_{\mathrm{W}}(-4)$ | Day -4 relative to symptom onset | 0.05 | (0.05, 0.05) | (0.01, 0.12) | (0.01, 0.12) |
| $a_{\mathrm{W}}(-3)$ | Day -3 relative to symptom onset | 0.06 | (0.06, 0.06) | (0.01, 0.13) | (0.01, 0.13) |
| $a_{\mathrm{W}}(-2)$ | Day -2 relative to symptom onset | 0.14 | (0.14, 0.14) | (0.01, 0.15) | (0.01, 0.15) |
| $a_{\mathrm{W}}(-1)$ | Day -1 relative to symptom onset | 0.07 | (0.07, 0.07) | (0.02, 0.17) | (0.01, 0.17) |
| $a_{\mathrm{W}}(0)$ | Day 0 relative to symptom onset | 0.16 | (0.16, 0.16) | (0.02, 0.18) | (0.01, 0.18) |
| $a_{\mathrm{W}}(1)$ | Day 1 relative to symptom onset | 0.17 | (0.17, 0.17) | (0.01, 0.17) | (0.01, 0.17) |
| $a_{\mathrm{W}}(2)$ | Day 2 relative to symptom onset | 0.02 | (0.02, 0.02) | (0.02, 0.16) | (0.01, 0.16) |
| $a_{\mathrm{W}}(3)$ | Day 3 relative to symptom onset | 0.10 | (0.10, 0.11) | (0.02, 0.15) | (0.01, 0.15) |
| $a_{\mathrm{W}}(4)$ | Day 4 relative to symptom onset | 0.12 | (0.12, 0.12) | (0.01, 0.14) | (0.01, 0.14) |
| $a_{\mathrm{W}}(5)$ | Day 5 relative to symptom onset | 0.08 | (0.08, 0.08) | (0.01, 0.14) | (0.01, 0.14) |
| $a_{\mathrm{W}}(6)$ | Day 6 relative to symptom onset | 0.07 | (0.07, 0.07) | (0.01, 0.13) | (0.01, 0.13) |
| $a_{\mathrm{W}}(7)$ | Day 7 relative to symptom onset | 0.05 | (0.05, 0.05) | (0.01, 0.12) | (0.01, 0.13) |
| $a_{\mathrm{W}}(8)$ | Day 8 relative to symptom onset | 0.05 | (0.05, 0.05) | (0.01, 0.13) | (0.01, 0.13) |
| $a_{\mathrm{W}}(9)$ | Day 9 relative to symptom onset | 0.08 | (0.08, 0.08) | (0.01, 0.14) | (0.01, 0.14) |
| $a_{\mathrm{W}}(10)$ | Day 10 relative to symptom onset | 0.14 | (0.14, 0.14) | (0.01, 0.16) | (0.01, 0.16) |

**Table S7.** Notation list of the mathematical symbols of the day-specific secondary attack rates $a_{\mathrm{C}}(t)$ in the health care layer for day $t$ relative to symptom onset. Italic small letters denote variables. Roman font indicates physical quantities. Estimate is the value yielded by the best firefly and used in Monte Carlo simulations and analysis, while range is the range of values among the set of best fireflies. The initial is the range of initial guesses in the initial population of 100 fireflies, and the constraints are limits enforced.

| Notation | Description | Estimate | Range | Initial | Constraints |
|---|---|---|---|---|---|
| $a_{\mathrm{C}}(-14)$ | Day -14 relative to symptom onset | 0.011 | (0.011, 0.011) | (0.000, 0.023) | (0.000, 0.023) |
| $a_{\mathrm{C}}(-13)$ | Day -13 relative to symptom onset | 0.006 | (0.006, 0.007) | (0.001, 0.018) | (0.001, 0.019) |
| $a_{\mathrm{C}}(-12)$ | Day -12 relative to symptom onset | 0.008 | (0.008, 0.009) | (0.001, 0.016) | (0.001, 0.016) |
| $a_{\mathrm{C}}(-11)$ | Day -11 relative to symptom onset | 0.005 | (0.005, 0.006) | (0.001, 0.018) | (0.001, 0.018) |
| $a_{\mathrm{C}}(-10)$ | Day -10 relative to symptom onset | 0.012 | (0.012, 0.012) | (0.001, 0.020) | (0.001, 0.020) |
| $a_{\mathrm{C}}(-9)$ | Day -9 relative to symptom onset | 0.015 | (0.015, 0.015) | (0.001, 0.018) | (0.001, 0.019) |
| $a_{\mathrm{C}}(-8)$ | Day -8 relative to symptom onset | 0.009 | (0.009, 0.009) | (0.001, 0.015) | (0.001, 0.015) |
| $a_{\mathrm{C}}(-7)$ | Day -7 relative to symptom onset | 0.002 | (0.002, 0.002) | (0.001, 0.012) | (0.001, 0.012) |
| $a_{\mathrm{C}}(-6)$ | Day -6 relative to symptom onset | 0.006 | (0.006, 0.006) | (0.001, 0.011) | (0.001, 0.011) |
| $a_{\mathrm{C}}(-5)$ | Day -5 relative to symptom onset | 0.003 | (0.003, 0.003) | (0.001, 0.011) | (0.001, 0.011) |
| $a_{\mathrm{C}}(-4)$ | Day -4 relative to symptom onset | 0.009 | (0.009, 0.009) | (0.001, 0.012) | (0.001, 0.012) |
| $a_{\mathrm{C}}(-3)$ | Day -3 relative to symptom onset | 0.009 | (0.009, 0.009) | (0.001, 0.014) | (0.001, 0.014) |
| $a_{\mathrm{C}}(-2)$ | Day -2 relative to symptom onset | 0.010 | (0.010, 0.010) | (0.001, 0.016) | (0.001, 0.016) |
| $a_{\mathrm{C}}(-1)$ | Day -1 relative to symptom onset | 0.009 | (0.009, 0.009) | (0.001, 0.018) | (0.001, 0.018) |
| $a_{\mathrm{C}}(0)$ | Day 0 relative to symptom onset | 0.014 | (0.014, 0.014) | (0.002, 0.018) | (0.001, 0.019) |
| $a_{\mathrm{C}}(1)$ | Day 1 relative to symptom onset | 0.016 | (0.016, 0.016) | (0.001, 0.018) | (0.001, 0.018) |
| $a_{\mathrm{C}}(2)$ | Day 2 relative to symptom onset | 0.015 | (0.015, 0.015) | (0.002, 0.017) | (0.001, 0.017) |
| $a_{\mathrm{C}}(3)$ | Day 3 relative to symptom onset | 0.010 | (0.010, 0.010) | (0.001, 0.015) | (0.001, 0.016) |
| $a_{\mathrm{C}}(4)$ | Day 4 relative to symptom onset | 0.013 | (0.013, 0.013) | (0.002, 0.014) | (0.001, 0.015) |
| $a_{\mathrm{C}}(5)$ | Day 5 relative to symptom onset | 0.004 | (0.004, 0.004) | (0.001, 0.014) | (0.001, 0.015) |
| $a_{\mathrm{C}}(6)$ | Day 6 relative to symptom onset | 0.005 | (0.005, 0.005) | (0.001, 0.014) | (0.001, 0.014) |
| $a_{\mathrm{C}}(7)$ | Day 7 relative to symptom onset | 0.006 | (0.006, 0.006) | (0.001, 0.013) | (0.001, 0.013) |
| $a_{\mathrm{C}}(8)$ | Day 8 relative to symptom onset | 0.001 | (0.001, 0.002) | (0.001, 0.014) | (0.001, 0.014) |
| $a_{\mathrm{C}}(9)$ | Day 9 relative to symptom onset | 0.004 | (0.004, 0.004) | (0.001, 0.015) | (0.001, 0.015) |
| $a_{\mathrm{C}}(10)$ | Day 10 relative to symptom onset | 0.016 | (0.016, 0.016) | (0.001, 0.017) | (0.001, 0.017) |

**Table S8.** Notation list of the mathematical symbols of the day-specific secondary attack rates $a_{\mathrm{M}}(t)$ in the municipality layer for day $t$ relative to symptom onset. Italic small letters denote variables. Roman font indicates physical quantities. Estimate is the value yielded by the best firefly and used in Monte Carlo simulations and analysis, while range is the range of values among the set of best fireflies. The initial is the range of initial guesses in the initial population of 100 fireflies, and the constraints are limits enforced.

| Notation | Description | Estimate | Range | Initial | Constraints |
|---|---|---|---|---|---|
| $a_{\mathrm{M}}(-14)$ | Day -14 relative to symptom onset | 0.012 | (0.012, 0.012) | (0.001, 0.014) | (0.000, 0.015) |
| $a_{\mathrm{M}}(-13)$ | Day -13 relative to symptom onset | 0.007 | (0.007, 0.007) | (0.001, 0.012) | (0.001, 0.012) |
| $a_{\mathrm{M}}(-12)$ | Day -12 relative to symptom onset | 0.002 | (0.002, 0.002) | (0.001, 0.010) | (0.001, 0.010) |
| $a_{\mathrm{M}}(-11)$ | Day -11 relative to symptom onset | 0.009 | (0.009, 0.009) | (0.001, 0.011) | (0.001, 0.011) |
| $a_{\mathrm{M}}(-10)$ | Day -10 relative to symptom onset | 0.011 | (0.011, 0.011) | (0.001, 0.013) | (0.001, 0.013) |
| $a_{\mathrm{M}}(-9)$ | Day -9 relative to symptom onset | 0.003 | (0.003, 0.003) | (0.001, 0.012) | (0.001, 0.012) |
| $a_{\mathrm{M}}(-8)$ | Day -8 relative to symptom onset | 0.005 | (0.005, 0.005) | (0.001, 0.009) | (0.001, 0.010) |
| $a_{\mathrm{M}}(-7)$ | Day -7 relative to symptom onset | 0.001 | (0.001, 0.001) | (0.001, 0.008) | (0.001, 0.008) |
| $a_{\mathrm{M}}(-6)$ | Day -6 relative to symptom onset | 0.005 | (0.005, 0.005) | (0.001, 0.007) | (0.001, 0.007) |
| $a_{\mathrm{M}}(-5)$ | Day -5 relative to symptom onset | 0.007 | (0.007, 0.007) | (0.001, 0.007) | (0.001, 0.007) |
| $a_{\mathrm{M}}(-4)$ | Day -4 relative to symptom onset | 0.006 | (0.006, 0.006) | (0.001, 0.008) | (0.001, 0.008) |
| $a_{\mathrm{M}}(-3)$ | Day -3 relative to symptom onset | 0.004 | (0.004, 0.004) | (0.001, 0.009) | (0.001, 0.009) |
| $a_{\mathrm{M}}(-2)$ | Day -2 relative to symptom onset | 0.002 | (0.002, 0.002) | (0.002, 0.010) | (0.001, 0.010) |
| $a_{\mathrm{M}}(-1)$ | Day -1 relative to symptom onset | 0.008 | (0.008, 0.008) | (0.001, 0.011) | (0.001, 0.011) |
| $a_{\mathrm{M}}(0)$ | Day 0 relative to symptom onset | 0.004 | (0.004, 0.004) | (0.001, 0.012) | (0.001, 0.012) |
| $a_{\mathrm{M}}(1)$ | Day 1 relative to symptom onset | 0.009 | (0.009, 0.009) | (0.002, 0.011) | (0.001, 0.011) |
| $a_{\mathrm{M}}(2)$ | Day 2 relative to symptom onset | 0.005 | (0.005, 0.005) | (0.001, 0.011) | (0.001, 0.011) |
| $a_{\mathrm{M}}(3)$ | Day 3 relative to symptom onset | 0.006 | (0.006, 0.006) | (0.001, 0.010) | (0.001, 0.010) |
| $a_{\mathrm{M}}(4)$ | Day 4 relative to symptom onset | 0.006 | (0.006, 0.006) | (0.001, 0.010) | (0.001, 0.010) |
| $a_{\mathrm{M}}(5)$ | Day 5 relative to symptom onset | 0.009 | (0.009, 0.009) | (0.001, 0.009) | (0.001, 0.009) |
| $a_{\mathrm{M}}(6)$ | Day 6 relative to symptom onset | 0.003 | (0.003, 0.003) | (0.001, 0.008) | (0.001, 0.009) |
| $a_{\mathrm{M}}(7)$ | Day 7 relative to symptom onset | 0.007 | (0.007, 0.007) | (0.001, 0.008) | (0.001, 0.008) |
| $a_{\mathrm{M}}(8)$ | Day 8 relative to symptom onset | 0.006 | (0.006, 0.006) | (0.001, 0.009) | (0.001, 0.009) |
| $a_{\mathrm{M}}(9)$ | Day 9 relative to symptom onset | 0.007 | (0.007, 0.007) | (0.001, 0.009) | (0.001, 0.010) |
| $a_{\mathrm{M}}(10)$ | Day 10 relative to symptom onset | 0.010 | (0.010, 0.010) | (0.001, 0.011) | (0.001, 0.011) |

**Table S9.** Notation list of the mathematical symbols of the state transition probabilities. Italic small letters denote variables. Roman font indicates physical quantities. Estimate is the value yielded by the best firefly and used in Monte Carlo simulations and analysis, while range is the range of values among the set of best fireflies. The initial is the range of initial guesses in the initial population of 100 fireflies, and the constraints are limits enforced.

| Notation | Description | Estimate | Range | Initial | Constraints |
|---|---|---|---|---|---|
| $p_{\mathrm{I^AR}}$ | Asymptomatic to recovered transition probability | 0.11 | (0.11, 0.11) | (0.06, 0.12) | (0.06, 0.12) |
| $p_{\mathrm{I^SR}}$ | Symptom onset to recovered transition probability | 0.58 | (0.58, 0.58) | (0.58, 1.00) | (0.57, 1.00) |
| $p_{\mathrm{I^CR}}$ | Critically ill to recovered transition probability | 0.59 | (0.59, 0.59) | (0.57, 0.98) | (0.56, 1.00) |

#### Area Between Curves Analysis

We calculated the expected time in a state before transition to the other for all state transition curves shown in main text Fig 5 using a Riemann sum approximation of the area under the curves. This is expressed in days in Table S10. Similarly, we quantified the discrepancy between the observed and simulated mean state transition between curves shown in Table S10. Positive values imply that the simulation curve lies above the observed curve (overestimation), while negative values indicate the opposite (underestimation). The magnitude represents the degree of disparity between observed and simulated curves, with larger absolute values indicating greater divergence. This measurement is such that if a simulation curve were to pass linearly through the center of the observed curve, the positive and negative areas would cancel each other out. By quantifying the total area between curves, we obtain a single metric that captures both the average distance between curves and shape differences between distributions. The disparity in the $I^A$ to R transition (-3.69) should be interpreted cautiously due to the extremely small sample size (n=2) in the observed data.

**Table S10.** Comparison between CovSyn simulation and Taiwanese COVID-19 observed data showing area differences between Kaplan-Meier curves and areas under individual curves for each state transition. Positive values in the Area Between Curves indicate CovSyn overestimation; negative values indicate underestimation.

| State Transition | Area Between Curves | Area under CovSyn Simulation (days) | Area under Taiwan Observed Data (days) |
|---|---|---|---|
| $I^A$ to $I^S$ | -1.08 | 6.14 | 7.22 |
| $I^A$ to C | 1.03 | 13.03 | 12.00 |
| $I^A$ to R | -3.69 | 49.31 | 53.00 |
| $I^S$ to $I^C$ | 3.50 | 11.70 | 8.20 |
| $I^S$ to C | -1.14 | 7.90 | 9.04 |
| $I^S$ to R | 4.75 | 33.31 | 28.56 |
| $I^C$ to R | 9.45 | 46.67 | 37.22 |
| $I^C$ to D | -0.62 | 19.96 | 20.58 |

#### Validation of individual-level data

In here, we provide the zoomed-out version of Fig 6 in the main text. Fig S1 covers the whole uncertainty range of the secondary attack rate.

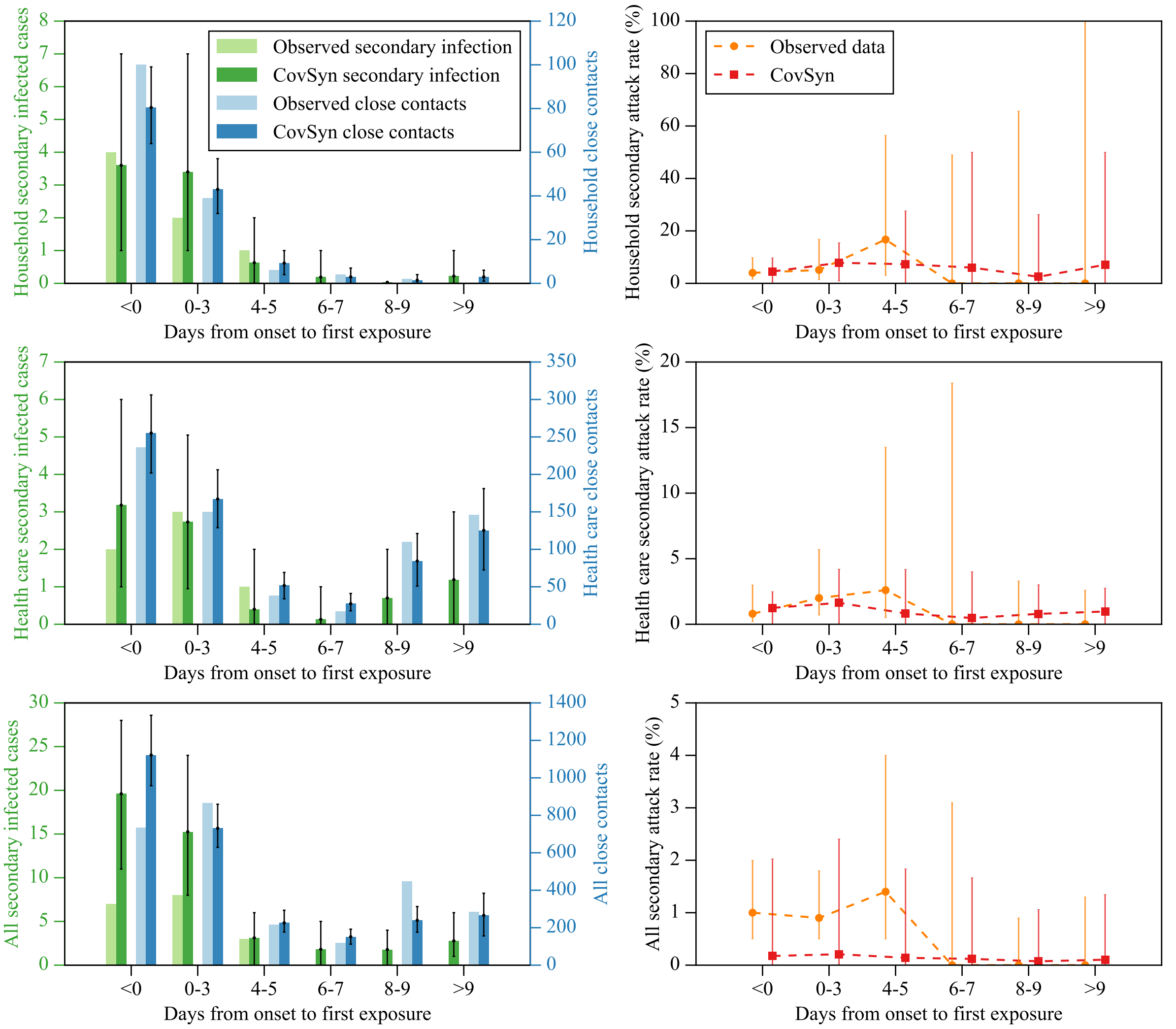


**Fig S1.** Comparison between CovSyn simulated data and observations from Cheng et al. 2020 across different contact layers: household (top), health care (middle), and all contacts (bottom). This plot is the zoomed-out version of Fig 6 in the main text. Left panels show secondary infected cases and close contacts. Right panels display the secondary attack rate, defined as the ratio of secondary infected cases to close contacts, expressed as a percentage. CovSyn was run for 100 Monte Carlo simulations, each with 100 source cases, to generate the 95% confidence intervals. The comparison demonstrates that our synthetic data exhibit the same trend in each layer.

### Validation of population-level data

The number of daily cases is so low the stochasticity dominants, while a substantial number of cases is preferable for a statistically robust comparison. For better visualization of the differences between our model simulations and actual observations, we utilized cumulative sums, which provide a clearer picture of overall model performance by not being dominated by noise.

Traditional error metrics such as Normalized Absolute Error (NAE) and Mean Absolute Percentage Error (MAPE) proved unsuitable for this analysis due to the monotonically increasing nature of cumulative cases. With these metrics, identical absolute errors yield substantially different NAE and MAPE values at different time points. Moreover, daily comparisons using these metrics were also not suitable due to the division-by-zero issues when actual case counts were zero. The full version of the Goodness of Fit (GOF) metrics table is provided in Table S11. The Mean Absolute Error (MAE), Mean Squared Error (MSE), Root Mean Squared Error (RMSE), and coefficient of determination ($R^2$) are presented with 95% confidence intervals.

Apart from the evaluation in the main text, in this section, we also include the Weighted Interval Score (WIS) to assess the fit between synthetic and real data, with results illustrated in Fig S2. For local confirmed cases, the WIS initially increased around 2020-01-07, reaching a first peak value of 86 on 2020-01-28, followed by a second peak of 131 on 2020-02-14, before eventually converging to 38. For local deaths, the WIS peaked at 40 on 2020-02-15 and 2020-02-16, subsequently converging to 11. These patterns indicate that CovSyn exhibited larger discrepancies during the early phase of the outbreak (2020-01-07 to 2020-02-16) before the substantial increase in cases began.

### Model structure

We show the extended version of the branching process model in Fig S3. The branching process can be easily mapped to daily cumulative infected, symptomatic, confirmed, critically ill, dead, and immune cases.

### Data synthesis

#### Draw demographic data

We gathered Taiwan demographic data including population pyramid, student rate, unemployment rate, part-time job rate, and full-time job rate with its occupation type. The population pyramid data for

**Table S11.** Comprehensive Goodness of Fit (GOF) metrics comparing Taiwanese population-level COVID-19 data with CovSyn synthetic data outputs. The metrics include Mean Absolute Error (MAE), Mean Squared Error (MSE), Root Mean Squared Error (RMSE), and coefficient of determination ($R^2$). Subscript c denotes cumulative values. MA refers to centered Moving Average applied to the time series. All metrics are presented with 95% confidence intervals.

| | **MAE** | **MAE$_\mathbf{c}$** | **MSE** | **MSE$_\mathbf{c}$** | **RMSE** | **RMSE$_\mathbf{c}$** | $R^2$ | $R^2_\mathbf{c}$ |
|---|---|---|---|---|---|---|---|---|
| Daily Confirmed | 0.4 | 6.3 | 0.8 | 73.0 | 0.9 | 7.6 | -0.4 | 0.9 |
| | (0.4, 0.5) | (2.2, 13.0) | (0.6, 1.1) | (9.7, 250.9) | (0.8, 1.0) | (3.1, 15.8) | (-1.0, -0.1) | (0.5, 1.0) |
| Daily Deaths | 0.1 | 1.5 | 0.1 | 6.3 | 0.3 | 2.3 | -2.7 | -2.8 |
| | (0.0, 0.1) | (0.6, 3.1) | (0.0, 0.1) | (0.7, 23.4) | (0.2, 0.4) | (0.9, 4.8) | (-5.8, -1.0) | (-12.9, 0.6) |
| 7-day MA Confirmed | 0.3 | 6.2 | 0.2 | 71.8 | 0.5 | 7.5 | 0.1 | 0.9 |
| | (0.2, 0.3) | (2.1, 12.9) | (0.1, 0.3) | (8.6, 249.8) | (0.4, 0.6) | (2.9, 15.8) | (-0.4, 0.5) | (0.5, 1.0) |
| 7-day MA Deaths | 0.1 | 1.5 | 0.0 | 6.3 | 0.1 | 2.3 | -4.5 | -2.8 |
| | (0.0, 0.1) | (0.6, 3.1) | (0.0, 0.0) | (0.7, 23.2) | (0.1, 0.2) | (0.8, 4.8) | (-11.2, -0.9) | (-13.0, 0.6) |
| 31-day MA Confirmed | 0.1 | 5.7 | 0.0 | 65.4 | 0.2 | 6.9 | 0.7 | 0.9 |
| | (0.1, 0.2) | (1.4, 12.8) | (0.0, 0.1) | (3.4, 244.4) | (0.1, 0.3) | (1.8, 15.6) | (0.4, 0.9) | (0.5, 1.0) |
| 31-day MA Deaths | 0.1 | 1.5 | 0.0 | 5.8 | 0.1 | 2.2 | -8.8 | -2.7 |
| | (0.0, 0.1) | (0.6, 3.0) | (0.0, 0.0) | (0.5, 22.1) | (0.0, 0.1) | (0.7, 4.7) | (-29.5, -0.7) | (-13.0, 0.7) |

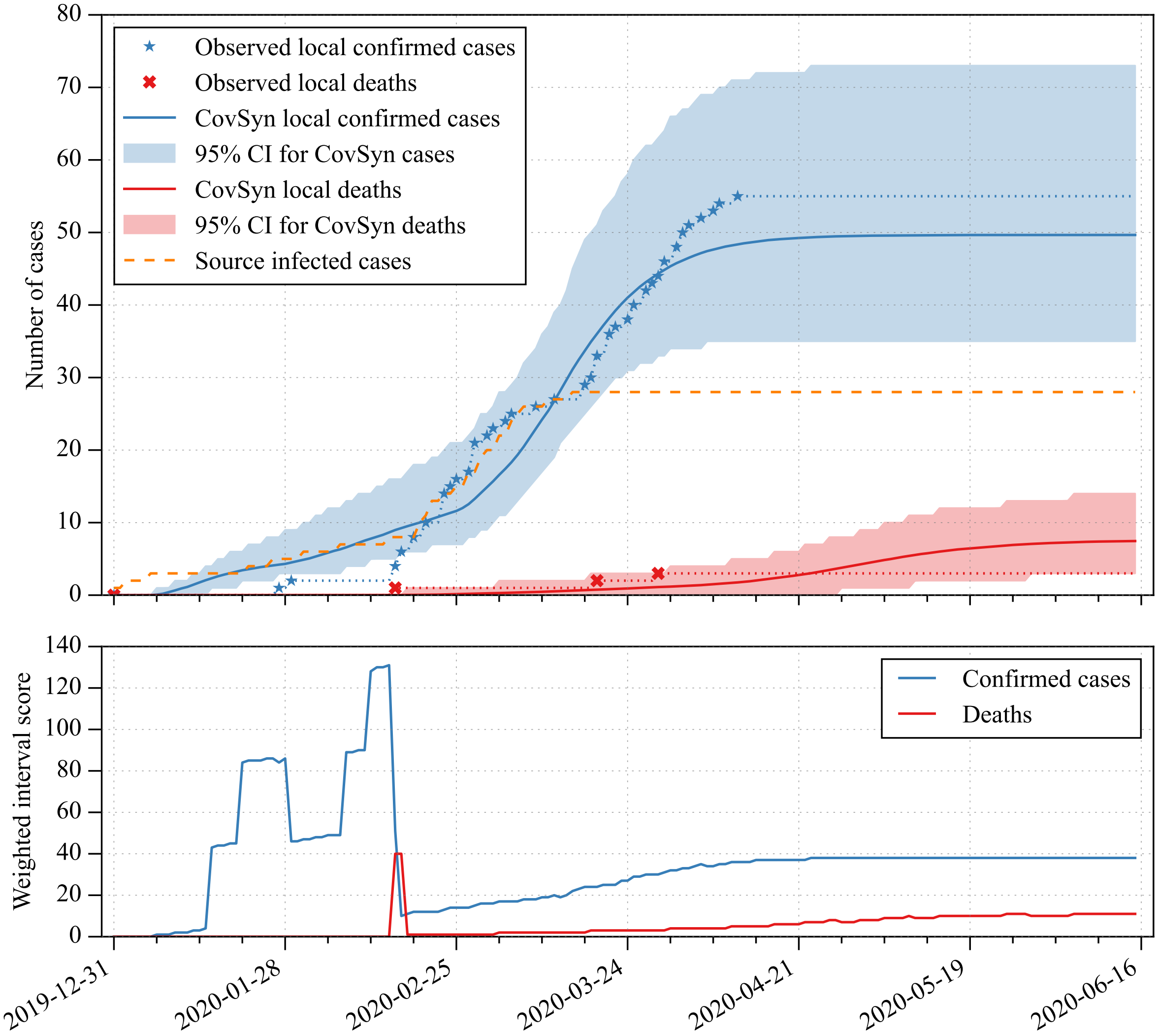


**Fig S2.** Comparison of observed Taiwan COVID-19 population-level data with CovSyn simulations. This plot is the extended version of Fig 7 in the main text. The top row shows the cumulative local confirmed cases and deaths. Blue stars and red dots show observed local cases and deaths, respectively. Dashed orange line indicates the 28 seed infections. Blue/red solid lines represent mean predictions from 1,000 Monte Carlo simulations, with 95% confidence intervals shown as shaded areas. The bottom row illustrates the Weighted Interval Score (WIS) comparison between observed and synthetic data.

2022 contained the population size from ages 0 to 99 and 'above 100'. We simplified the age 'above 100' to just 100 years old.

The student rate data contained student numbers from elementary school to university (ages 7 to 22). The 5th grade or above university student rate was ignored due to the low number of students compared to the other grade students.

For the unemployment rate, part-time job, and full-time job rate data, they were originally grouped as age $15-19, 20-24, \ldots, 60-64$. We assumed the rate in each age group to be constant, and padded

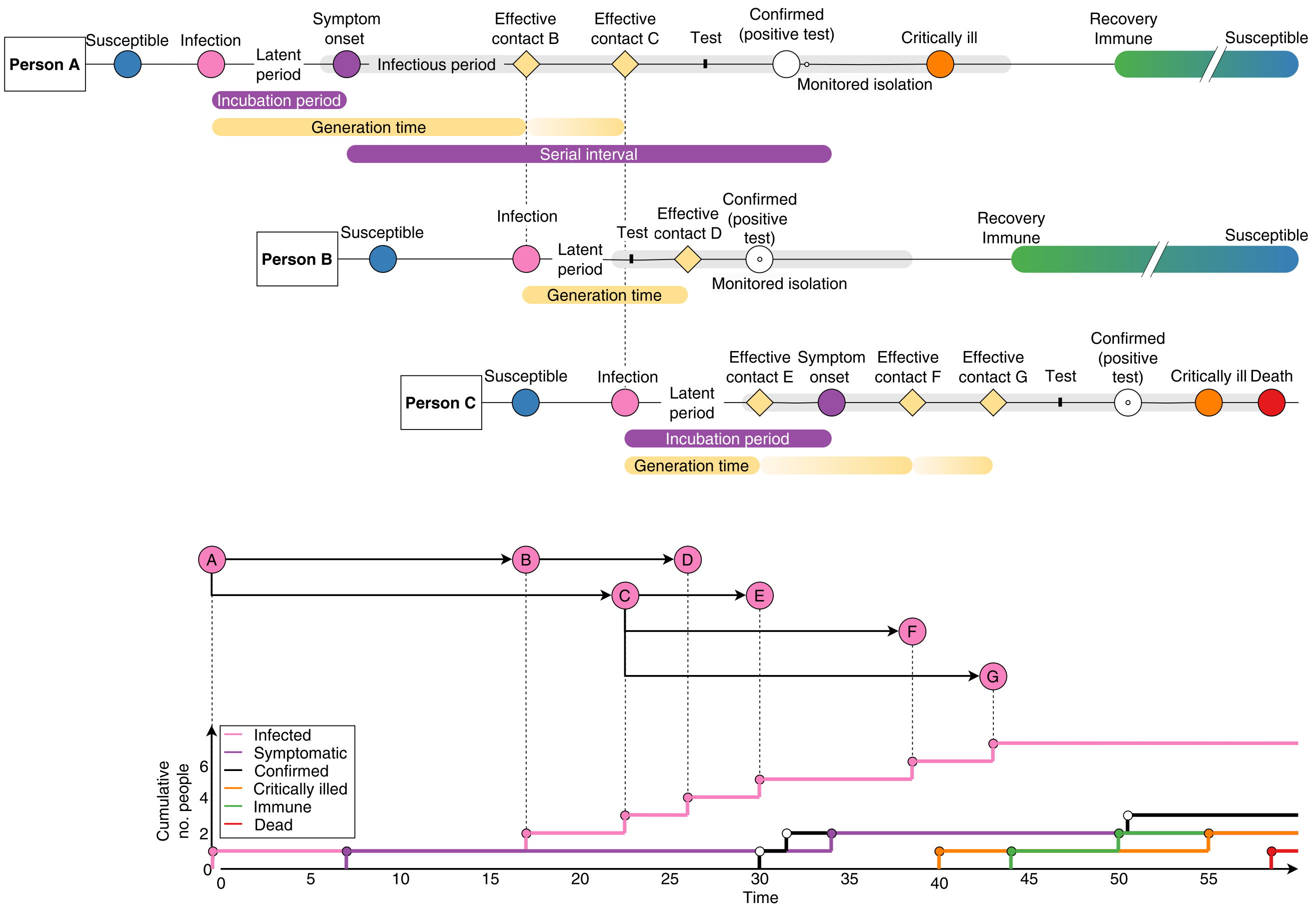


**Fig S3.** Illustration of the simulated course of disease branching process and mapping to daily cumulative infected, symptomatic, confirmed, critically ill, dead, and immune cases. This plot is the extended version of Fig 8 in the main text. Case B and C are the effective contacts of case A, case D get infected by case B, and case E to G are infected by case C. The bottom plot shows that the resulting chains can be mapped into the cumulative cases per day, which follow an exponential growth trend.

zeros for ages below 15 and above 65.

For the occupation synthesis, we need to determine the student status, employment status, and occupation. If the individual is not a student and unemployed, the occupation field will remain empty. If the individual is a student and employed, the occupation will be selected from the part-time job set, otherwise it will be selected from the full-time job set.

The pseudocode for the demographic data synthesis is shown in Algorithm S1.

#### Draw social data

Next, we randomly sampled the contact size for five different layers, which were household, school class, work group, clinic, and municipality. The Taiwanese data contained household size for each couty and was grouped with the family size from 1 to 5 and above 6. School class size data was described in the previous section. For the work group layers, the original data grouped the company size for different occupations as $< 5, 5-9, 10-19, 20-29, 30-39, 40-49, 50-99, 100-199, 200-299, 300-499, 500-999$, and above 1000. For each occupation, we calculated the probability that a person can be in each sized company. We assumed that the probability in each size group was identical. To calculate the specific size of a company size above 1000, we split the probability of 1000 to be a certain number of equal pieces so that the probability of each piece will be less than the probability of a company size 999.

The pseudocode for the social context data synthesis is shown in Algorithm S2.

#### Draw contact data

##### Contact network

The daily contacts of each infection case (marked by preceding I) were synthesized and stored as matrices. Fig S4 shows an example of the time series of contact of the first cluster containing 2 infected cases (I1 and I2). The pink nodes represent the infected cases at the time of infection. The effective contact is marked as a yellow diamond on day 2 in this example. As only contacts that appear in the infectious period (shaded grey area) of the source case are at risk of infection, we synthesized contacts only after infection and before monitored isolation. The start time of infectious period was defined as the minimum date of (i) confirmed date, (ii) date of a secondary infection, or (iii) symptom onset date. Day 0 was defined as the day the first source case I1 was infected. The smaller size nodes (1 to 7) were the index cases of contacts but not infected. On day 1, I1 had contact with index 1 to 5. On day 2, index 5 was updated to be I2 after the case turns out to be a confirmed case. On day 3, I2 turned out to be infectious and contacted with indexes 4 and 7. The same index case might be in contact with source case during multiple days (*e.g.* case 4 has contacted I1 on days 1, 3, and 6 and has contacted I2 on days 3 and day 5).

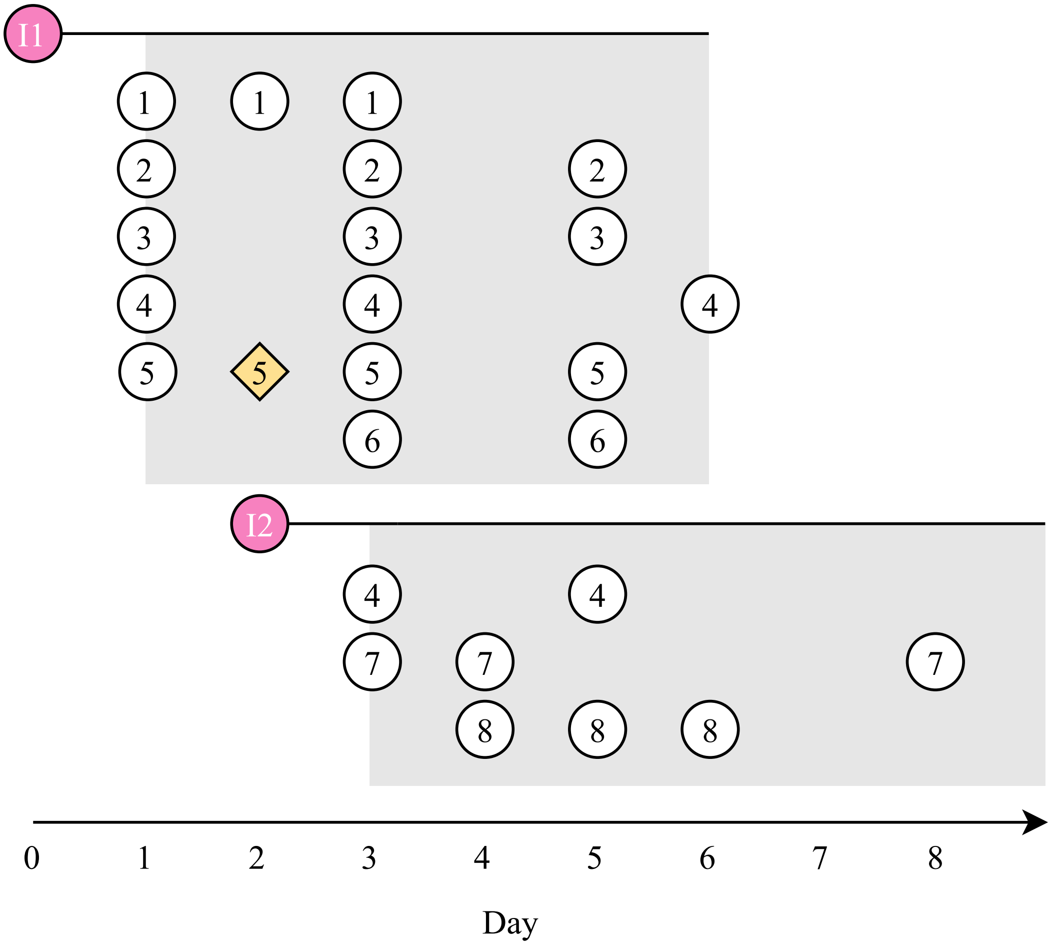


**Fig S4.** An illustration of the daily contacts of cases I1 and I2. The pink nodes represent the infected cases at the time of infection. The infectious period is represented as a shaded grey area and the nodes inside the area are contacts with their indices. The yellow diamond is the effective contact.

##### Effective contact rate

Transmission of infection required susceptible individuals, infected individuals and effective contact between them. The effective contact rate ($\beta$) was the effective contacts in a given population per time unit. It can be expressed as the multiplication of the total contact rate (the total number of contacts per unit time, $\gamma$) and the transmission risk ($\rho$).

$$\beta = \gamma \times \rho \tag{1}$$

The transmission risk (or secondary attack rate, $\rho$) is the probability of the effective transition of the disease (at a specific time) such that $\rho \in [0, 1]$ and the direct assessment of it is,

$$\rho = \frac{x}{n}, \tag{2}$$

where $x$ is the number of secondary infected cases and $n$ is the number of contacts in a group, [1]. This also implies that the effective contact rate will always be less or equal to the total contact rate since not all the contacted individuals would be infected.

**Calculate adjusted daily secondary attack rate**

We calculated the daily secondary attack rate based on estimates from Ge *et al.* [2]. Their study showed that the daily secondary attack rate ranged from -14 to 10 days from the symptom onset date to the exposure date. To implement the secondary attack rate for cases with different infectious periods and incubation times, we stretched or compressed the daily secondary attack rate based on how long the infectious period was. Consider the incubation period (symptom onset date) as the centre, if the time from symptom onset to the end of the infectious period was different from 10 days, we stretched or compressed the daily secondary attack rate from Ge *et al.* to be the same length as the simulated case. We did the same thing for the 14 days before the symptom onset data. If the case was an asymptomatic case, which meant there was no time of symptom onset, we resized the secondary attack rate array from -14 days to end of the infectious period.

Each layer of contacts would have a different secondary attack rate. The mean value of the secondary attack rate for household, school, workplace, and health care was 10.1, 2.3, 3.4, and 0.4 [2–4] The secondary attack rate for the municipality layer was set to 0.2 since we assumed it is smaller than that of any other layer. We normalised the daily secondary attack rate to fit the mean value for each layer. The secondary attack rate vector was adjusted by considering the overdispersion rate of the source case and the age of the secondary contact (Algorithm S7). We further optimized the secondary attack rates by the Firefly algorithm. An illustration of the resulting daily contacts and the daily secondary attack rates for each layer are shown in Fig S5.

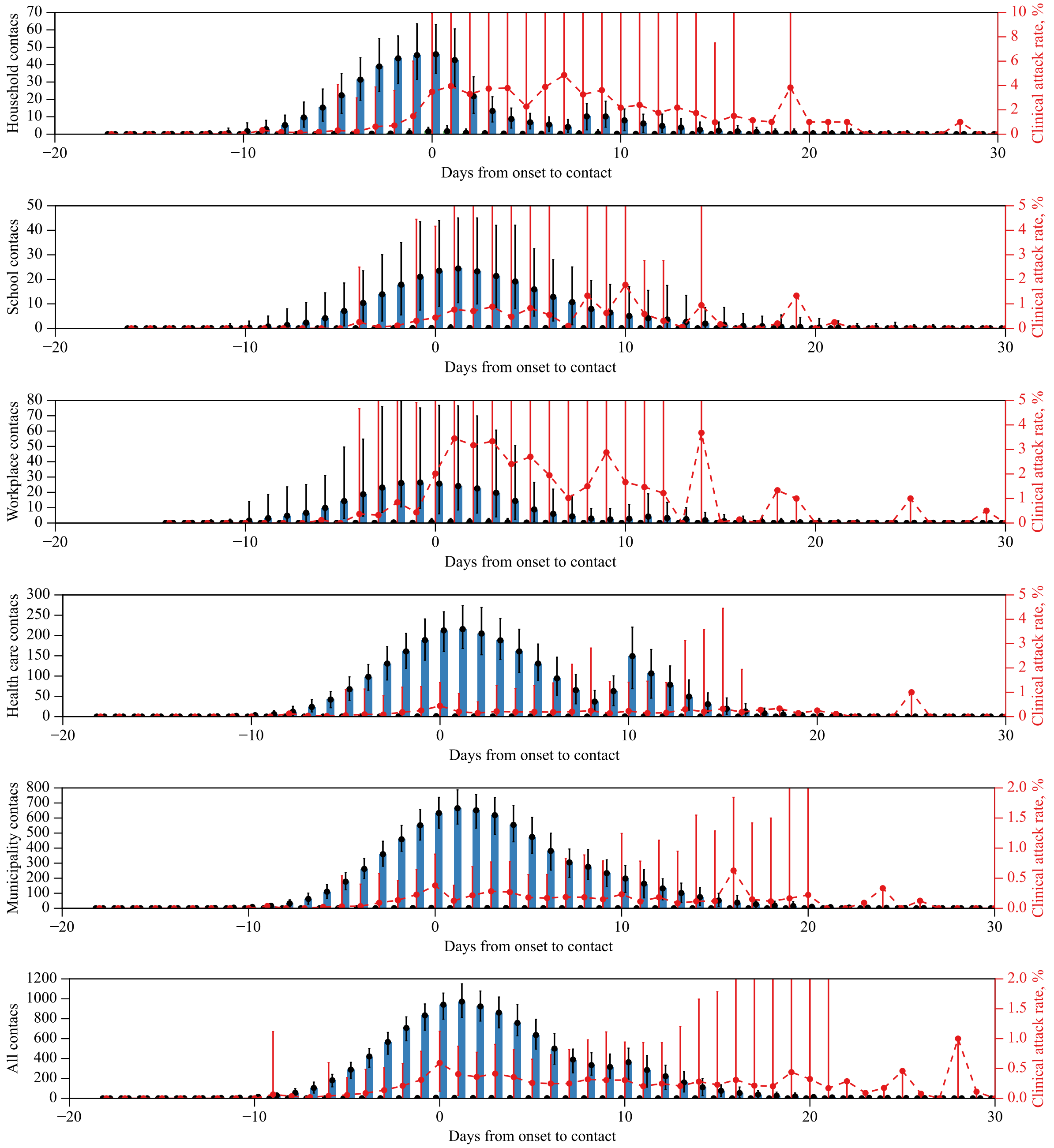


**Fig S5.** Contact day vs infection day for 100 source cases. The contact day is counted starting from symptom onset of the source case, i.e., day 0. Blue, green, and red colors represent the number of contacts, the number of infections, and the secondary attack rate, respectively. The 95% confidence interval is shown.

##### Comparison to reported values

We here show the comparison between our synthetic data's commonly used epidemiological parameters and the reported values in Fig S6. Since our algorithm does not specifically simulate the infectious period of asymptomatic cases, the resulting mean value is similar to the infectious period of symptomatic cases, which is overestimated compared to the reported values.

#### Algorithm

**Algorithm S1** Draw demographic data

**Input:** age probabilities, gender probabilities by age, student probabilities by age and gender, employment probabilities by age and gender, job probabilities by gender and employment type
**Output:** age, gender, and job

age ← randomly select an age with weights from age probabilities
gender ← randomly select male or female with weights from gender probabilities by age
student_status ← randomly determine student status based on student probabilities by age and gender
employment_status ← randomly determine employment status based on employment probabilities by age and gender
job ← randomly select job based on student_status, employment_status, and job probabilities by gender and employment type

**Algorithm S2** Draw social context data

**Input:** age, job, municipality data, family size distributions by municipality, school class size probabilities by age, workplace size probabilities by job, hospital probabilities, hospital size probabilities by hospital
**Output:** household size $n_{\mathrm{H}}$, school class size $n_{\mathrm{S}}$, work group size $n_{\mathrm{W}}$, clinic size $n_{\mathrm{C}}$, municipality size $n_{\mathrm{M}}$

municipality ← randomly select a municipality with weights from municipality data
$n_{\mathrm{M}} \leftarrow$ population of the selected municipality
$n_{\mathrm{H}} \leftarrow$ randomly select household size using family size distribution by municipality
$n_{\mathrm{S}} \leftarrow$ randomly select school class size based on age and school class size probabilities by age
$n_{\mathrm{W}} \leftarrow$ randomly select work group size based on job and workplace size probabilities by job
hospital ← randomly select a hospital based on hospital probabilities
$n_{\mathrm{C}} \leftarrow$ randomly determine clinic size based on hospital size probabilities by hospital

**Algorithm S3** Draw vaccination status

**Input:** Vaccine efficacy $e_{\mathrm{V}} \in \mathbb{R}_{[0,1]}$, vaccination rate $r_{\mathrm{V}} \in \mathbb{R}_{[0,1]}$
**Output:** Vaccination status $s_{\mathrm{V}} \in \{\mathrm{True}, \mathrm{False}\}$

$s_{\mathrm{V}} \leftarrow$ random choice True or False with success rate $r_{\mathrm{V}} \times e_{\mathrm{V}}$

To draw the contact data each day, we first need to simulate the number of contacts for the current individual. Then, we proceed with assigning the contact day for each contact. This typically requires iteratively performing random sampling for each day with the corresponding daily contact probability.

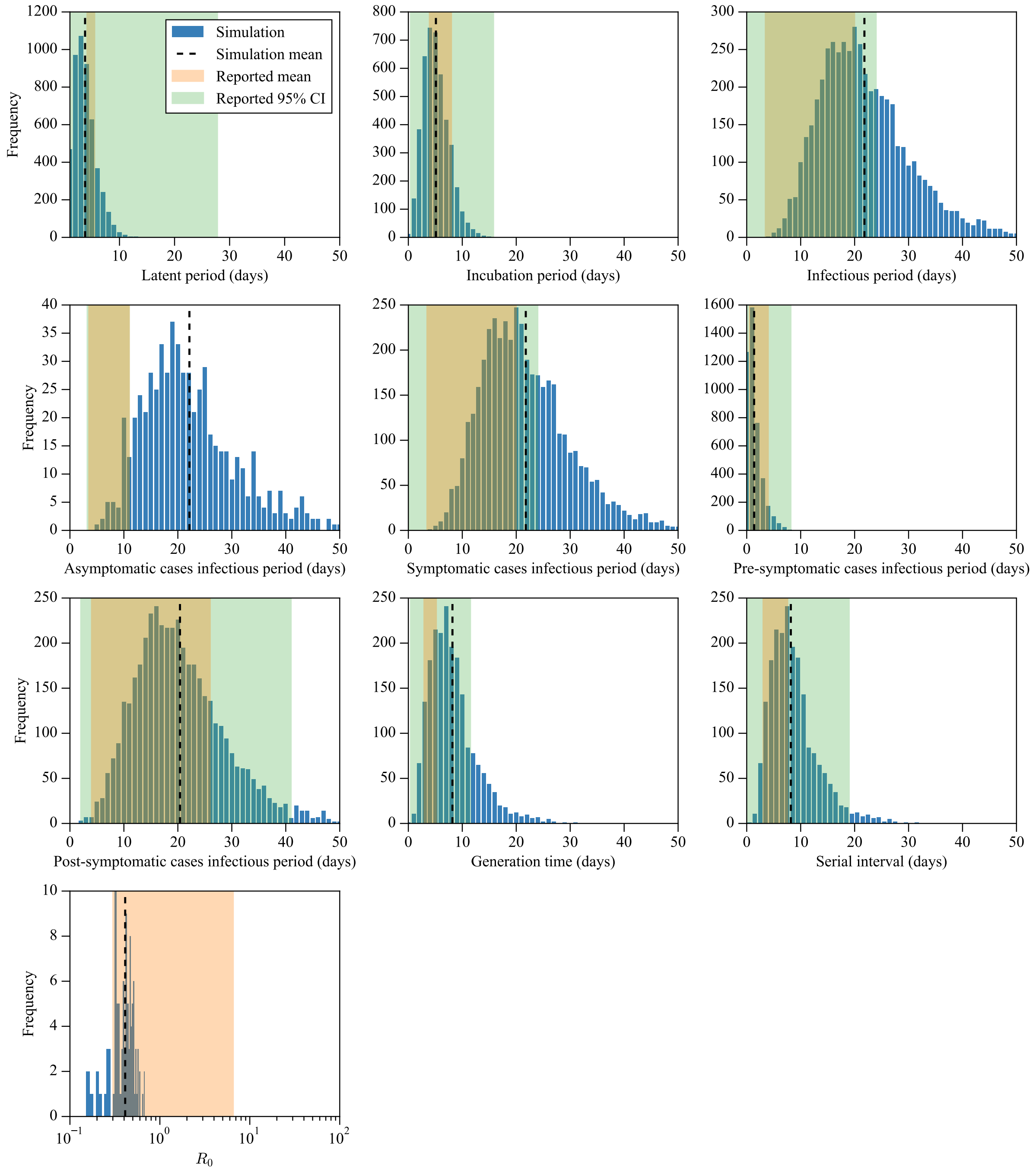


**Fig S6.** Statistics of the synthetic data generated by our model. The basic reproduction number ($R_0$) is calculated by averaging the number of effective contacts for each infected case. This plot is the extended version of Fig 3 in the main text. The simulation results are depicted as blue bars with mean values shown as black dashed lines. We also report the range of reported statistics from multiple sources. The range of reported mean values is plotted in orange shaded areas, and the range of the 95% confidence interval is plotted in green shaded areas. The reported latent period is based on [4, 5], incubation period on [4, 6, 7], infectious period on [8], generation time on [9–11], serial interval on [7, 11], and $R_0$ on [12–20]

**Algorithm S4** Determine overdispersion state

**Input:** Overdispersion rate $r_{\mathrm{O}} \in \mathbb{R}_{[0,1]}$
**Output:** Overdispersion state $s_{\mathrm{O}} \in \{\mathrm{True}, \mathrm{False}\}$

$s_{\mathrm{O}} \leftarrow$ random choice True or False with success rate $r_{\mathrm{O}}$

**Algorithm S5** Calculate daily secondary attack rate

**Input:** Daily secondary attack rate vector $\boldsymbol{a}_i \in \mathbb{N}_{[0,1]}$, where $i$ is the social layers $\in \{\mathrm{H}, \mathrm{S}, \mathrm{W}, \mathrm{C}, \mathrm{M}\}$, infectious period $\tau_{\mathrm{F}} \in \mathbb{N}_0$, incubation period $\tau_{\mathrm{I}} \in \mathbb{N}_0$, latent period $\tau_{\mathrm{L}} \in \mathbb{N}_0$, time of beginning monitored isolation $t_{\mathrm{M}} \in \mathbb{N}_0$
**Output:** Adjusted daily secondary attack rate vectors $\tilde{\boldsymbol{a}}_i \in \mathbb{N}_{\{0,1\}}$, $i \in \{\mathrm{H}, \mathrm{S}, \mathrm{W}, \mathrm{C}, \mathrm{M}\}$

**for** $i \in \{\mathrm{H}, \mathrm{S}, \mathrm{W}, \mathrm{C}, \mathrm{M}\}$ **do**
  **if** symptomatic case **then**
    $\tilde{\boldsymbol{a}}_i \leftarrow$ stretch $\boldsymbol{a}_i$ to length $\tau_{\mathrm{F}}$ with center index at $\tau_{\mathrm{I}}$
  **else**
    /* Asymptomatic case */
    $\tilde{\boldsymbol{a}}_i \leftarrow$ stretch $\boldsymbol{a}_i$ to length $\tau_{\mathrm{F}}$ with center index at 0
  **end if**
  $\tilde{\boldsymbol{a}}_i \leftarrow$ pad zero vector with length $\tau_{\mathrm{L}}$ in front
  **if** $t_{\mathrm{M}} \geq \tau_{\mathrm{L}} + \tau_{\mathrm{F}}$ **then**
    $\tilde{\boldsymbol{a}}_i \leftarrow$ append zero vector with length $t_{\mathrm{M}} - (\tau_{\mathrm{L}} + \tau_{\mathrm{F}})$ in the end
  **else**
    Set the final $(\tau_{\mathrm{L}} + \tau_{\mathrm{F}}) - t_{\mathrm{M}}$ elements in $\tilde{\boldsymbol{a}}_i$ to zeros
  **end if**
**end for**

**Algorithm S6** Draw from previously infected set

**Input:** Previously infected set $\mathbb{I}$, population set $\mathbb{P}$
**Output:** Previously infected status $y \in \{\mathrm{True}, \mathrm{False}\}$

$y \leftarrow$ random choice True or False with success rate $len(\mathbb{I})/len(\mathbb{P})$

**Algorithm S7** Draw infection status and effective contact day vector

**Input:** Adjusted daily secondary attack rate vector $\tilde{\boldsymbol{a}}_i \in \mathbb{R}_{[0,1]}$, contact day vector $\boldsymbol{m} \in \mathbb{N}_{\{0,1\}}$, natural immunity status $s_{\mathrm{N}} \in \{\mathrm{True}, \mathrm{False}\}$, vaccination status $s_{\mathrm{V}} \in \{\mathrm{True}, \mathrm{False}\}$, overdispersion rate $r_{\mathrm{O}} \in \mathbb{R}_{[0,1]}$, overdispersion weight $w_{\mathrm{O}} \in \mathbb{R}_{[0,1]}$, age of the secondary contact $g_{\mathrm{c}} \in \mathbb{N}_0$

**Output:** Infection status $s_{\mathrm{I}} \in \{\mathrm{True}, \mathrm{False}\}$, effective contact day vector $\boldsymbol{m}_{\mathrm{e}} \in \mathbb{N}_{\{0,1\}}$

Initialize $\boldsymbol{m}_{\mathrm{e}}$ to be zero vector with length of $\tilde{\boldsymbol{a}}_i$<br>
$s_{\mathrm{I}} \leftarrow False$<br>
**for** $j \in \{1, \ldots, length(\boldsymbol{m})\}$ **do**<br>
    **if** $\boldsymbol{m}[j] == 1$ **then**<br>
        **if** $s_{\mathrm{N}} == False$ **&** $s_{\mathrm{V}} == False$ **then**<br>
            $\tilde{\boldsymbol{a}}_i^{\mathrm{a}}[j] \leftarrow$ calculate the age adjusted daily secondary attack rate /* Algorithm S8 */<br>
            $s_{\mathrm{O}} \leftarrow$ determine the overdispersion state /* Algorithm S4 */<br>
            **if** $s_{\mathrm{O}} == True$ **then**<br>
                $x \leftarrow$ random choice True or False with success rate $\tilde{\boldsymbol{a}}_i^{\mathrm{a}}[j] \times w_{\mathrm{O}}$<br>
            **else**<br>
                $x \leftarrow$ random choice True or False with success rate $\tilde{\boldsymbol{a}}_i^{\mathrm{a}}[j]$<br>
            **end if**<br>
            **if** $x == True$ **then**<br>
                $\boldsymbol{m}_{\mathrm{e}}[j] \leftarrow 1$<br>
                $s_{\mathrm{I}} \leftarrow True$<br>
                Break<br>
            **end if**<br>
        **end if**<br>
    **end if**<br>
**end for**

**Algorithm S8** Calculate age adjusted daily secondary attack rate

**Input:** Adjusted daily secondary attack rate vector $\tilde{\boldsymbol{a}}_i \in \mathbb{R}_{[0,1]}$, age of the secondary contact $g_{\mathrm{c}} \in \mathbb{N}_{[0,100]}$, age related risk ratio vector of the secondary attack rate $\boldsymbol{r}_a$

**Output:** Age adjusted daily secondary attack rate $\tilde{\boldsymbol{a}}^{\mathrm{a}}$

$\tilde{\boldsymbol{a}}^{\mathrm{a}} \leftarrow \tilde{\boldsymbol{a}}_i \times \boldsymbol{r}_a[g_{\mathrm{c}}]$<br>
$\tilde{\boldsymbol{a}}^{\mathrm{a}}[\tilde{\boldsymbol{a}}^{\mathrm{a}} > 1] = 1$

To ensure each contact has at least one contact day, a straightforward approach would be to apply a while loop until at least one element in the contact vector becomes true. However, this approach is not efficient since it might be trapped in an infinite loop when the daily contact probabilities are too small. Therefore, instead of using a while loop, we first simulate a contact matrix with the first contact day. The pseudo-code is shown in Algorithm S11. This algorithm implements two phases of random sampling. In the first phase, it uses a simple for loop to go through each day and randomly assigns a contact day based on the daily probabilities. If these probabilities are too small, some contacts might miss the chance of assignment and result in a row of all false values. In such cases, we consider the relative probability of each day, *i.e.*, we normalize the daily contact probabilities to sum to 1. We then apply these normalized probabilities to sample a single contact day, ensuring that every contact is assigned exactly one day of interaction. After the first contact matrix, we can apply our vectorized algorithm to draw the contact data for each day, shown in Algorithm S10.

**Algorithm S9** Draw contacts each day

**Input:** Probability of contact $p$, consecutive contact probability $p_{\mathrm{c}}$, contact probability when healthy $p_{\mathrm{h}}$, contact probability when symptomatic $p_{\mathrm{s}}$, steepness of the logistic contact probability function $s$, phase relative to symptom onset for symptomatic (days) $t_{\mathrm{PS}}$, phase relative to symptom onset for recovering to normal (days) $t_{\mathrm{PN}}$, overdispersion rate $r_{\mathrm{O}}$, and overdispersion weight $w_{\mathrm{O}}$ for each social layer $\{\mathrm{H}, \mathrm{S}, \mathrm{W}, \mathrm{C}, \mathrm{M}\}$ (household, school, workplace, health care, municipality); social data object with social context sizes

**Output:** Contact matrix for all the social layers $\boldsymbol{M}$

1: **for** each social layer $i \in \{\mathrm{H}, \mathrm{S}, \mathrm{W}, \mathrm{C}, \mathrm{M}\}$ **do**
2: Extract social size $n_i$ from social data object
3: $\boldsymbol{M}_i \leftarrow$ draw social contacts each day /* Algorithm S10 */
4: **end for**
5: $\boldsymbol{M} \leftarrow \{\boldsymbol{M}_{\mathrm{H}}, \boldsymbol{M}_{\mathrm{S}}, \boldsymbol{M}_{\mathrm{W}}, \boldsymbol{M}_{\mathrm{C}}, \boldsymbol{M}_{\mathrm{M}}\}$
6: **return** $\boldsymbol{M}$

**Algorithm S10** Draw social contacts each day

**Input:** For the source case in the social layer i $\in \{\mathrm{H}, \mathrm{S}, \mathrm{W}, \mathrm{C}, \mathrm{M}\}$, time of symptom onset $t_{\mathrm{I^S}} \in \mathbb{N}_0$, incubation period $\tau_{\mathrm{I}} \in \mathbb{N}_0$, period from infection to monitored isolation $\tau_{\mathrm{M}} \in \mathbb{N}_0$, and social context size $n_{\mathrm{i}} \in \mathbb{N}_0$; probability of contact $p \in \mathbb{R}_{[0,1]}$, consecutive contact probability $p_{\mathrm{c}} \in \mathbb{R}_{[0,1]}$, contact probability when healthy $p_{\mathrm{h}} \in \mathbb{R}_{[0,1]}$, contact probability when symptomatic $p_{\mathrm{s}} \in \mathbb{R}_{[0,1]}$, steepness of the logistic contact probability function $s \in \mathbb{R}_+$, phase relative to symptom onset for symptomatic (days) $t_{\mathrm{PS}} \in \mathbb{R}_+$, phase relative to symptom onset for recovering to normal (days) $t_{\mathrm{PN}} \in \mathbb{R}_+$.

**Output:** A contact matrix, $M \in \{0,1\}^{c \times (\tau_{\mathrm{M}}+1)}$, specifying the days of contact

$c \leftarrow B(n_i, p)$ /* Sample number of contacts from Binomial distribution */
**if** $c = 0$ **then**
  **return** empty matrix
**end if**
$\boldsymbol{p} \leftarrow$ zero vector with length $\tau_{\mathrm{M}} + 1$ /* Daily contact probability vector */
**if** $\tau_{\mathrm{I}} = \texttt{NaN}$ **then**
  /* Asymptomatic case */
  $\boldsymbol{p} \leftarrow ones(\tau_{\mathrm{M}} + 1) \times p_{\mathrm{h}}$
**else**
  /* Symptomatic case */
  $\boldsymbol{t} \leftarrow$ vector from 0 to $\tau_{\mathrm{M}}$
  $\boldsymbol{p} \leftarrow f_{p_L}(\boldsymbol{t} - t_{\mathrm{I^S}}, p_{\mathrm{h}}, p_{\mathrm{s}}, s, t_{\mathrm{PS}}, t_{\mathrm{PN}})$
**end if**
$M \leftarrow$ generate the first contact matrix /* Algorithm S11 */
**for** $j \in \{1, \ldots, \tau_{\mathrm{M}}\}$ **do**
  /* Simulate subsequent contacts */
  $\mathrm{mask}_{\mathrm{no}} \leftarrow \{\text{indices where } M_{:,j} = 0\}$ /* No contact indices */
  prev $\leftarrow M_{:,j-1}$ /* Previous day's contacts */
  **for** $i \in \{1, \ldots, c\}$ **do**
    **if** $i \in \mathrm{mask}_{\mathrm{no}}$ **then**
      /* No contact yet today */
      **if** $\mathrm{prev}_i = 1$ **then**
        $M_{i,j} \leftarrow B(1, p_{\mathrm{c}})$ /* Consecutive contact probability */
      **else**
        $M_{i,j} \leftarrow B(1, \boldsymbol{p}_j)$ /* Base probability for day j */
      **end if**
    **end if**
  **end for**
**end for**
**return** $M$

**Algorithm S11** Generate first contact matrix

**Input:** Contacts number $c \in \mathbb{N}$, number of days $n_d \in \mathbb{N}$, daily contact probabilities $\boldsymbol{p} = \{p_1, \ldots, p_{n_d}\}$ where $p_i \in \mathbb{R}_{[0,1]}$

**Output:** A contact matrix $M \in \{0,1\}^{c \times n_d}$, where each row contains exactly one true value

$M \leftarrow$ zero matrix of size $c \times n_d$
$U \leftarrow \emptyset$ /* Set of unassigned contact indices */
**for** $i \in \{1, \ldots, c\}$ **do**
  /* Phase 1: Natural probability assignment */
  contact_assigned $\leftarrow$ false
  **for** $j \in \{1, \ldots, n_d\}$ **do**
    **if** random() $< p_j$ **then**
      $M_{i,j} \leftarrow true$
      contact_assigned $\leftarrow$ true
      **break**
    **end if**
  **end for**
  **if** not contact_assigned **then**
    $U \leftarrow U \cup \{i\}$ /* Add to unassigned set */
  **end if**
**end for**
**if** $U \neq \emptyset$ **then**
  /* Phase 2: Force assignment for remaining */
  $\boldsymbol{p_n} \leftarrow \boldsymbol{p} / \sum_{i=1}^{n_d} p_i$ /* Normalize probabilities */
  **for** $i \in U$ **do**
    $j \leftarrow$ random choice index from $\{1, \ldots, n_d\}$ with probabilities $\boldsymbol{p_n}$
    $M_{i,j} \leftarrow 1$
  **end for**
**end if**
**return** $M$
